# Detection of Neoplasms by Metagenomic Sequencing of Cerebrospinal Fluid

**DOI:** 10.1101/2021.05.13.21256918

**Authors:** Wei Gu, Andreas M. Rauschecker, Elaine Hsu, Kelsey C. Zorn, Yasemin Sucu, Scot Federman, Allan Gopez, Shaun Arevalo, Hannah A. Sample, Eric Talevich, Eric D. Nguyen, Marc Gottschall, Carl A. Gold, Bruce A.C. Cree, Vanja Douglas, Megan B. Richie, Maulik P. Shah, S. Andrew Josephson, Jeffrey M. Gelfand, Steve Miller, Linlin Wang, Tarik Tihan, Joseph L. DeRisi, Charles Y. Chiu, Michael R. Wilson

## Abstract

**Importance:** Malignant neoplasms of the central nervous system (CNS) are frequently not detected by cerebrospinal fluid (CSF) flow cytometry or cytology, and clinical phenotypes can overlap with inflammatory meningoencephalitis.

**Objective:** To determine whether an existing CSF metagenomic next-generation sequencing (mNGS) assay can identify a hallmark of malignant neoplasms — aneuploidy — in difficult-to-diagnose cases of CNS malignancy.

**Design:** Two retrospective, case-control studies included a total of 155 samples from patients with an eventual diagnosis of a CNS malignancy (n=59 patients) and controls with other CNS diseases (n=73 patients). The first study was used to evaluate test performance in positive and negative controls. The second study was used to assess the potential utility of aneuploidy detection in patients whose CSF was sent for mNGS because of suspected neuroinflammatory disease who were ultimately found to have a CNS malignancy.

**Setting:** This is a single site study at a large tertiary care center, University of California San Francisco, that enrolled from 2014 to 2019.

**Participants:** The test performance case-control study enrolled positive control patients with a CNS malignancy (n=47 patients) and negative controls with other neurologic diseases (n=56 patients) who had had CSF flow cytometry and/or cytology performed. The second case-control study enrolled patients with suspected neuroinflammatory disease who were ultimately diagnosed with a CNS malignancy (n=12) and other neurologic disease controls (n=17).

**Main Outcome(s) and Measure(s):** The primary outcome measures were the performance characteristics of detecting aneuploidy in CSF by a cell-free DNA mNGS assay compared to cytology and/or flow cytometry and the tumor fraction in CSF from patients with CNS malignancies.

**Results:** Across the two case-control studies, the overall sensitivity of the CSF mNGS assay for detecting aneuploidy in patients ultimately diagnosed with a CNS malignancy was 75% (63-96%, 95% CI), and specificity was 100% (96-100%, 95% CI). Notably, CSF mNGS detected aneuploidy in 64% of the non-diagnostic cytology and flow cytometry cases in the test performance study and in 55% of the cases with suspected neuroinflammatory disease who were ultimately diagnosed with a CNS malignancy. Of the cases in whom aneuploidy was detected, 90% had multiple chromosomal copy number variants with tumor fractions ranging from 31% to 49%.

**Conclusions and Relevance:** Metagenomic NGS of CSF, originally designed to diagnose neurologic infections, detects evidence of CNS malignancies (i.e., aneuploidy) in cases where CSF flow cytometry and/or cytology were negative with a low risk of false positive results.

**3 Key Points:** *Question:* Can CSF metagenomic NGS, a test designed to diagnose infections, also detect genetic signatures of cancer in patients with suspected neuroinflammatory disease?

*Findings:* Across two case-control studies of patients with negative CSF cytology and/or flow cytometry, CSF mNGS detected genetic evidence for a malignancy with a sensitivity of 75% (63-85%, 95% CI) and specificity of 100% (96-100%, 95% CI).

*Meaning:* CSF mNGS, an assay with low sample volume requirements that does not require the preservation of cell integrity, has the potential to complement cancer detection by CSF flow cytometry and cytology.

## Introduction

Determining the etiology of meningoencephalitis is a challenge, with approximately half of cases going undiagnosed^1–3^. Major diagnostic considerations include occult infections, autoimmune disorders, and a variety of malignancies, some of which can manifest as leptomeningeal carcinomatosis^1^. However, malignant conditions are notoriously difficult to diagnose even after multiple large volume cerebrospinal fluid (CSF) exams. This difficulty may be due to the paucicellular nature of CSF, including small numbers of malignant cells or the need to preserve the cells’ morphology before they lyse. Since CNS malignancies, infections, and inflammatory syndromes can clinically and radiologically overlap, a single test that can detect more than one type of hard-to-diagnose condition would have potential clinical utility.

To detect CNS infections, our group and others have reported on the utility of metagenomic next-generation sequencing (mNGS) for detecting a wide array of pathogens in CSF^4–12^. mNGS generates sequences from all the genetic material in a sample, and the vast majority of the sequences in CSF are human. Previously, we and others have found CNS tumor DNA in CSF^13–17^. To detect CNS malignant neoplasms, we hypothesized that CSF mNGS human data could be repurposed to detect aneuploidy as a specific marker. Indeed, aneuploidy and other large copy number variations (CNVs) are prevalent in primary and metastatic CNS tumors, with ∼90% of all malignant tumors harboring aneuploid (abnormally numbered) chromosomes^18^. To test this hypothesis, we employed the depth of coverage method used in noninvasive prenatal testing to detect aneuploidy^19,20^.

We first characterized test performance on a pilot set of positive and negative control CSF samples obtained from patients undergoing clinical evaluation for a CNS malignancy, primarily for CNS lymphoma. Next, we performed the same analyses on samples from a nested case-control study of subjects enrolled in an ongoing, prospective study of patients with suspected neuroinflammatory disease^5,7^.

## Results

### Test Performance Study

#### Study Population

We identified 125 CSF samples from 103 patients (Figure 1B, Table 1, Table S1) from residual specimens in the clinical laboratory, including 55 samples from 47 patients with a CNS malignancy. Positive controls were from lumbar punctures (n=46), drains (n=4), shunts (n=1), and Ommaya reservoirs (n=4). Lymphomas comprised the majority of malignancies (36 of 55, 65% of cases). A subset of the positive controls were diagnosed by CSF cytology and/or flow cytometry (n=31 samples from 23 patients). The remainder of the patients with a CNS malignancy (n=24 samples from 24 patients) were negative or indeterminate on CSF cytology and/or flow cytometry but were eventually diagnosed with a malignancy using a combination of tissue biopsy (n=22/24), repeat CSF cytology and/or flow cytometry and Neuroradiology and Oncology diagnostic consensus with intention to treat. clinical follow-up with expert consensus.

**Figure 1:**
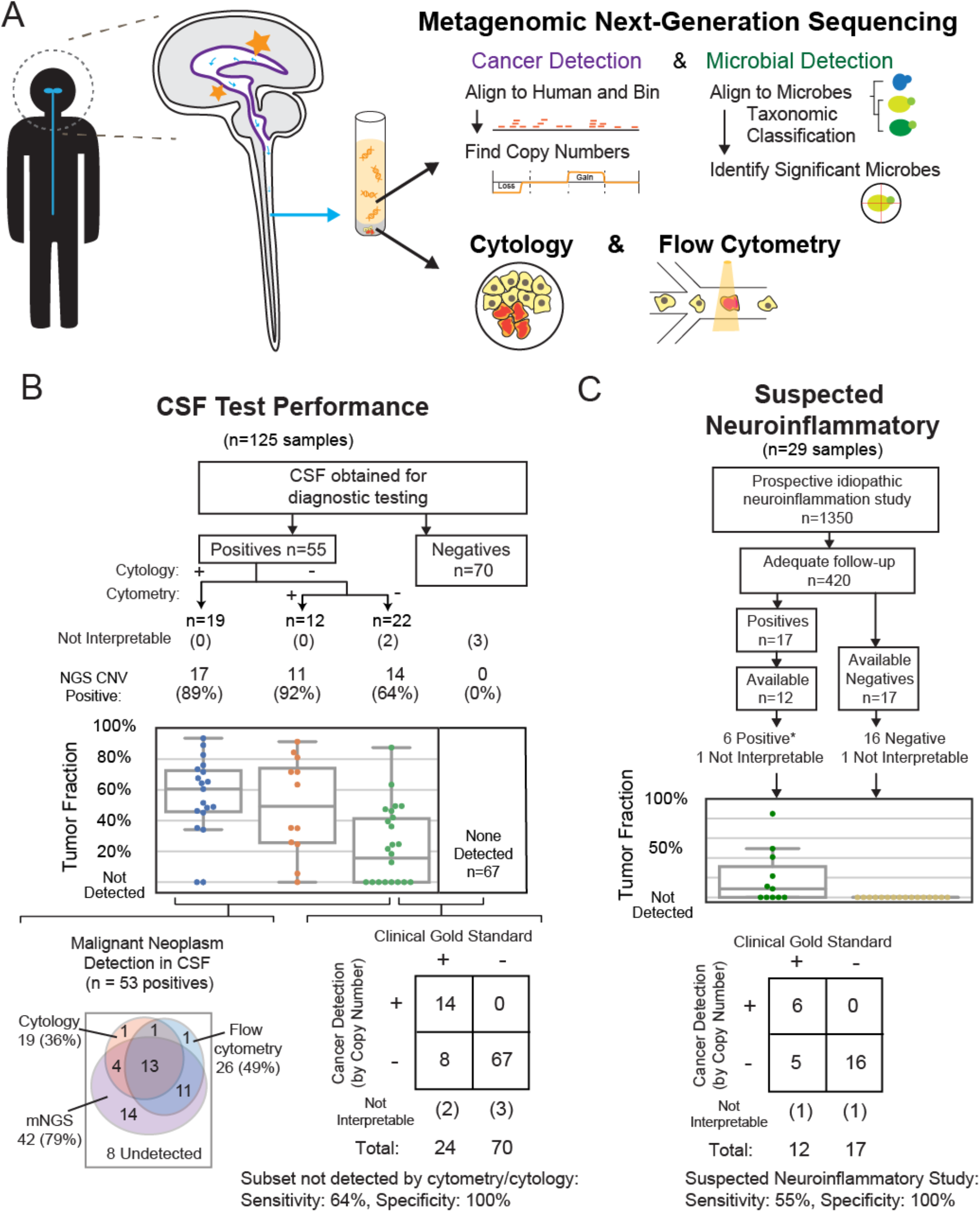
**(A) Schematic of the mNGS testing strategy**. Brain lesions release pathogen or cancer cell-free DNA into the cerebrospinal fluid space. Metagenomic next-generation sequencing was performed on all specimens to simultaneously assess for somatic aneuploidy or significant pathogens. **(B) Case-Control Study 1: CSF Test Performance**. Cases were enrolled from residual CSF referred for flow cytometry and/or cytology testing. Of the total 125 cases, 55 positives were diagnosed clinically with cancer and 70 were negative controls (ultimately diagnosed with an infectious or autoimmune condition). The cases were subcategorized by the ability of conventional testing to detect the malignancy or required orthogonal testing by later testing or more invasive procedures. The estimated tumor fractions shown are based on the amplitude of the copy change. Case details are found in Table S1. (Bottom Left) Venn diagrams of detection by cytology, flow cytometry, and mNGS. (Bottom Right) Contingency table using the positives that were missed by conventional testing (flow cytometry and/or cytology) of CSF. **(C) Case-Control Study 2: Suspected Neuroinflammatory Diseases**. Cases (n=12) and controls (n=17) from a prospective study of patients with suspected neuroinflammatory disease. Long term follow-up confirmed CNS tumors in positive cases. Negative controls were from patients eventually diagnosed with CNS autoimmune disorders. Case details are found in Table S2. (Bottom) Contingency table based on these positives and negatives.

Negative controls (n=70 samples from 56 patients) were required to have an alternative diagnosis (e.g., infection, autoimmune disease) that explained their neurologic presentation. Negative controls were from lumbar punctures (n=53), drains (n=12), and shunts (n=5). Infectious disease testing results for a subset of these subjects, including pathogens identified by mNGS, have been previously reported^12^. See Methods for further details.

#### Sequencing Results

The median depth of sequencing was 7 million reads (IQR 5.0-9.4 million reads). All but 4 specimens underwent paired-end sequencing of more than 125 base pairs on an Illumina sequencer. The remaining 4 specimens underwent single-end sequencing. Large (>10 Mbp) CNVs across the genome were characterized by the bioinformatic pipeline described in Methods and by blinded interpretation of the copy ratio plots. A minority of the results (4%: 2 positive controls, 3 negative controls) did not generate interpretable copy ratio plots, presumably due to low DNA input, and were not included in the following performance calculations.

The sensitivity for detecting at least one large CNV in all positive control samples was 79% (95% CI 66%-89%), excluding two cases that were not interpretable (Figure 1B). Specifically, in the positive controls with positive CSF cytology (n=19) or flow cytometry (n=12), sensitivity was 89% (95% CI 67%-99%) and 92% (95% CI 62%-100%), respectively. In the 22 positive controls where cytology and/or flow cytometry results were negative (benign) or inconclusive (e.g., atypical cells), 64% (95% CI 41-83%) were positive by mNGS for aneuploidy or other large CNVs above 10 Mbp (Figure 1B). In the 44 positive samples obtained by lumbar puncture rather than a drain or Ommaya reservoir, the sensitivity was 75% (95% CI 60-87%). The specificity was 100% (95% CI 95%-100%) in the negative controls (n=70) (i.e., no false positives were identified).

We calculated tumor fractions based on maximal deviations from the copy number baseline and assumptions about the tumor copy numbers in that region (see Methods). The estimated tumor fractions of all mNGS positives (n=55) was a median of 49% (IQR 35-72%). In the subset of positives where the traditional methods (cytology and flow cytometry) failed to detect the malignancy (n=24), the tumor fraction of mNGS positive cases was still surprisingly high at 41% (IQR 24-49%). Of the detectable cases, 90% had multiple CNVs, 81% had 4 or more CNVs, and 29% had 10 or more CNVs. Given that most cases were positive for lymphoma, we inspected the non-lymphoma cases (n=12), and the overall tumor fraction was also high at 48% (IQR 32-61%).

### Suspected Neuroinflammatory Disease Study

Once we were able to gather preliminary data on the ability of the mNGS assay to detect aneuploidy in positive and negative control samples, we tested its performance in a clinically relevant cases and controls retrospectively identified from a more extensive study enrolling patients with suspected neuroinflammatory diseases. These are patients who can be hard to differentiate clinically between infectious, autoimmune, and neoplastic conditions. Thus, a CSF mNGS assay might be ordered to look broadly for a neurologic infection. After we screened the first 1,350 patients enrolled between 2014 to 2019, 420 (31%) had adequate follow-up and an eventual clinical diagnosis. Of these, we identified 12 cases with remaining CSF that had a clinical diagnosis of either a primary CNS tumor (n= 9) or a systemic tumor that had metastasized to the CNS (n=3) as documented by prior sites of involvement (five additional cases lacked residual CSF). All CSF samples were obtained by lumbar puncture. CSF cytology and flow cytometry showed benign results in 10 cases and “atypical cells” in two. The cases were matched with 17 negative controls (autoimmune diagnosis without CNS malignancy) from the same study (Figure 1C, Table 2, Table S2).

CNVs were detected by CSF mNGS in 6 of the 12 cases with malignancy, and one case was not interpretable (presumably due to insufficient DNA input) for a sensitivity of 55% (23-83%, 95% CI) (Figure 1C, Table S2). Three of the cases with malignancy had more than 10 large (>10 Mbp) CNVs, and the remaining had 1, 8, and 8 CNVs. The median tumor fraction of the positives was 31% (14-47%, 95% CI). Similar to the first study, no CNVs were detected in the negative controls (n=16; 1 case was not interpretable, presumably due to insufficient DNA input) for a specificity of 100% (79-100% (95% CI)). All positive NGS findings from the nested case-control study are shown in Figure 2D. These cases include 2 cases of primary CNS lymphoma (PCNSL), 2 cases of intravascular lymphoma (IL), and 2 cases of metastatic melanoma.

**Figure 2:**
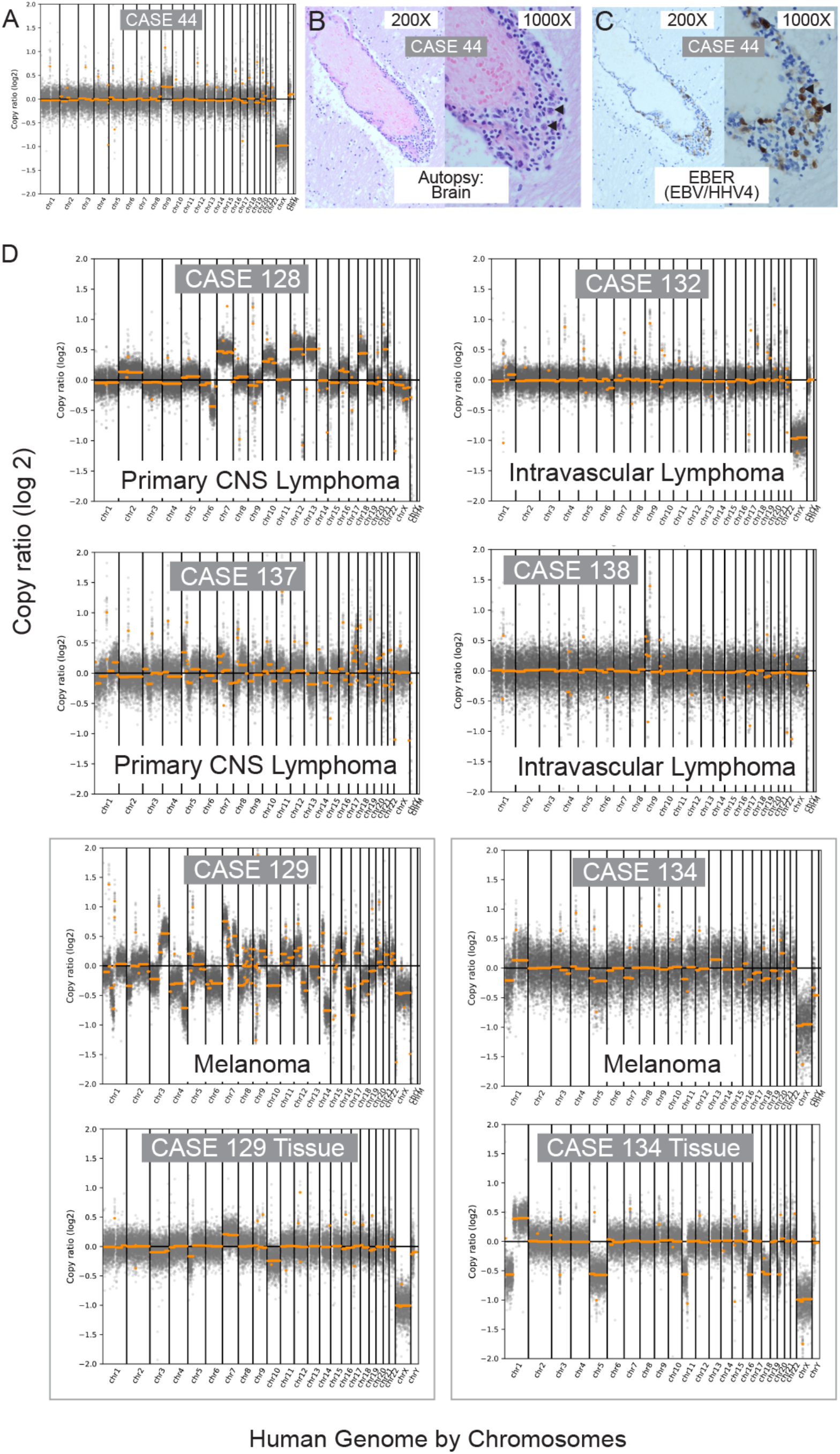
Results of Specific Undiagnosed Cases. A) Case 44: Metagenomic NGS of Case 44 shows Trisomy 9 at a tumor fraction of 36% and EBV positivity. Conventional methods by brain biopsy and cytology of the cerebrospinal fluid were nondiagnostic. Flow cytometry showed atypical cells and was not definitive. The diagnosis was confirmed by autopsy two weeks after the CSF sampling. B) Case 44: H&E of a post-mortem brain section. Arrowheads highlight large perivascular B cells that have large, irregular nuclei with loose chromatin. C) Case 44: EBER RNA *in situ* hybridization stain (brown) of a brain section from the autopsy. Arrowheads highlight the same set of large cells positive for EBER as evidence of EBV RNA expression in malignant lymphoma. D) The copy ratio plots of all 6 positive cases from the suspected neuroinflammatory disease case-control study. For Cases 129 and 134, the comparable copy ratio plot derived from directly sequencing the neoplasm is shown.

When combining all interpretable samples from both studies, the sensitivity of CNV detection by mNGS was 75% (63-85%, 95% CI), and specificity was 100% (96-100%, 95% CI). Given that some patients had multiple samples, and the overlap of two cases between the two studies, we recalculated an accuracy for unique patients across all studies by using the earliest sample. On a unique patient basis, the sensitivity was 71% (58-83%, 95% CI) and the specificity was 100% (95-100%, 95% CI).

### CSF CNV Characteristics Reflect CNVs Detected in Primary Tumor Tissue

Across both studies, there were 13 patients diagnosed with a CNS malignancy who had residual tumor tissue or a previous molecular or cytogenetic analysis performed. Of these 13 cases, 4 did not have CNVs detected by CSF mNGS. The remaining 9 cases demonstrated matching CNVs between the associated cancer tissue, although exact matches were precluded presumably by tumor genome evolution and subsequent genetic heterogeneity (Figure 2D, Supplemental Materials).

### Tumor Detection is not Limited to Radiographic Lesions that Abut the CSF Space

Brain MRIs from patients ultimately diagnosed with a CNS malignancy (64 of 66 cases over the two studies) were reviewed by a neuroradiologist blinded to the CSF mNGS CNV results (Figure 3). MRIs were within 2 days of CSF collection in 50% of cases and within 1 week in 80% of the cases. In cases for whom aneuploidy was detected by CSF mNGS and imaging was available, 80% (37 of 46) had either leptomeningeal involvement or were abutting the CSF space. However, the few remaining cases detected by mNGS either had lesions that did not abutt the CSF space (6 of 46) or did not have any radiographic lesions within 7 days of collection (3 of 46). This is in contrast to a prior smaller study where all five CNS tumors that did not abutt the CSF were not detected^14^.

**Figure 3.**
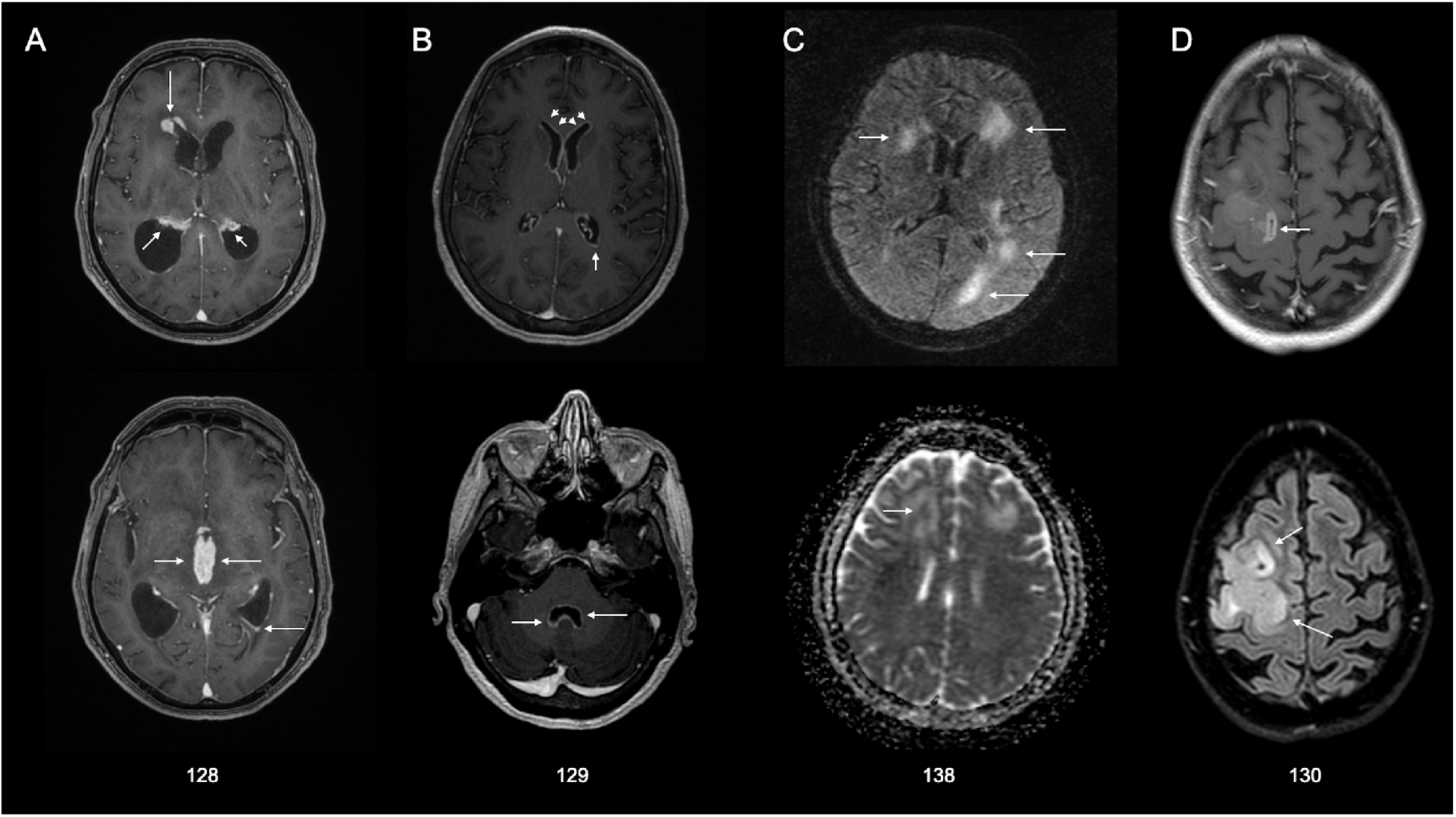
Imaging of abnormalities in four suspected neuroinflammatory cases at or near the time of brain biopsy. (A) In Case 128, post-contrast T1-weighted images demonstrate thick areas of enhancement along nearly all ependymal surfaces, including the lateral ventricles (top image), third ventricle (bottom image), and fourth ventricle (not shown), resulting in acute hydrocephalus. The imaging appearance was thought to be consistent with disseminated tuberculosis, less likely lymphoma due to limited restricted diffusion. The final diagnosis was primary CNS lymphoma. (B) In Case 129, post-contrast T1-weighted images demonstrate extensive, thin ependymal enhancement along the lateral ventricles (top image) and fourth ventricle (bottom image), with minimal adjacent signal abnormality in the parenchyma. The imaging appearance was thought most consistent with infectious ventriculitis, less likely carcinomatosis. The final diagnosis was melanoma. (C) In Case 138, motion-degraded T2/FLAIR (top image) and apparent diffusion coefficient (ADC) (bottom image) demonstrate asymmetric white matter signal abnormality with variable foci of mildly restricted diffusion. In this immunocompromised patient, images were thought be potentially consistent with progressive multifocal leukoencephalopathy. The final diagnosis was intravascular lymphoma. (D) In Case 130, post-contrast T1-weighted (top image) and T2/FLAIR (bottom image) sequences demonstrate expansile signal abnormality with areas of heterogeneous enhancement. Glioblastoma was diagnosed with brain biopsy. White arrows indicate areas of abnormalities.

### Case Vignettes

#### Case 44

Adult man with a history of HIV (CD4 count <35 cells/mm^3^) not on antiretroviral therapy presented with gait instability and was found to have multifocal (frontal and cerebellar) enhancing lesions on brain MRI primarily concerning for CNS lymphoma, toxoplasmosis, or tuberculosis. Despite extensive CSF testing, including cytology (benign), flow cytometry (“atypical B-cells that can be seen in either reactive or neoplastic conditions”), and a non-diagnostic brain biopsy (i.e., gliosis with mild chronic inflammation), a definitive diagnosis was not reached before he passed three weeks later. An autopsy confirmed a diagnosis of EBV-positive PCNSL (Figure 2B-C). We performed mNGS on the CSF aspirated from a drain over two weeks, and detected a trisomy 9 in all three samples with a tumor fraction ranging from 36 to 51% (Figure 2A). Epstein-Barr virus (EBV) was detected by CSF mNGS and plasma EBV PCR, and confirmed by EBV immunohistochemistry in the autopsy brain tissue.

#### Case 128

An adult man presented with 4 years of progressive cognitive decline and idiopathic hydrocephalus complicated by 1 week of worsening encephalopathy. Brain MRI showed multifocal thick mass-like ependymal and subependymal enhancement with extensive surrounding T2/FLAIR signal abnormality involving the deep and periventricular white matter (Figure 3A). An examination of his CSF obtained from a lumbar puncture revealed a white blood cell (WBC) count of 33/μL. CSF cytology documented atypical cells on three separate CSF samples, but a bone marrow biopsy and peripheral flow cytometry were negative for malignancy. The consensus diagnosis was CNS lymphoma, based on the atypical cells seen on cytology, the clinical presentation, the radiographic findings, and the initial, dramatic therapeutic response to steroids that unfortunately was followed by the development of septic shock and death one month after presentation. CSF mNGS detected a tumor fraction of 85% and over 10 chromosomal-level CNVs.

#### Case 129

An adult woman with a history of metastatic melanoma treated with surgery, radiation, and chemotherapy presented with 9 months of headache, 2 months of diplopia, nausea, and vomiting. She was treated empirically for bacterial and fungal meningitis after a CSF examination revealed a WBC count of 332/μL and a RBC count of 655/μL. MRI of the spinal cord showed smooth, diffuse leptomeningeal enhancement predominating in the lower thoracic cord and around the conus medullaris, favoring a non-malignant meningitis, with carcinomatosis considered less likely. Similarly, a brain MRI showed subependymal enhancement along the ventricular surfaces and the tentorium (Figure 3B). CSF cytology in two specimens was benign, and flow cytometry showed no evidence of a lymphoproliferative disorder. A brain biopsy diagnosed metastatic melanoma. CSF mNGS detected a tumor fraction of 41% with more than 20 CNVs, including 4 CNVs found in the patient’s lymph node biopsy in which melanoma had been found.

#### Case 132

An adult man with a history of diabetes presented with 7 months of asymmetrical weakness and was found to have longitudinally extensive transverse myelitis and enhancing brain lesions on MRI. He was initially empirically treated with glucocorticoids for a presumed inflammatory demyelinating condition. An examination of his CSF revealed a WBC count of 7/μL. Three separate CSF specimens sent for cytology were benign, and a bone marrow biopsy with flow cytometry showed no evidence of a lymphoproliferative disorder. Intravascular large B cell lymphoma was diagnosed by brain biopsy. CSF mNGS detected 3 CNVS and a tumor fraction of 8.7%.

#### Case 134

An adult man with a past medical history of cardiomyopathy status post defibrillator placement, chronic obstructive pulmonary disease, diabetes mellitus and 6 years of gait difficulty and communicating hydrocephalus requiring placement of an extraventricular drain who presented with two years of progressive short-term memory loss and worsening gait that prompted an evaluation for an underlying neurodegenerative process. A CSF examination unexpectedly revealed a WBC count of 168 cells/uL. CSF cytology showed atypical cells, but flow cytometry showed no evidence of a lymphoproliferative disorder. A CT scan of the brain and spinal cord showed possible leptomeningeal enhancement over the surface of the medulla and spinal cord, and a focus of nodular enhancement in the interhemispheric fissure abutting the left pregenual cingulate. The diagnosis of melanoma was ultimately made by brain biopsy. CSF mNGS detected a tumor fraction of 11% with more than 10 CNVs. Seven of the CSF mNGS CNVs overlapped with the CNVs detected in the patient’s brain biopsy.

#### Case 137

An adult man with a history of atrial fibrillation on anti-coagulation and treated syphilis who developed gait difficulty was found to have multifocal parenchymal lesions with ependymal involvement on brain MRI of unclear etiology. He had a dramatic benefit from empiric steroids after no diagnosis could be made with CSF testing (no safe brain biopsy target initially). Five months later, when the patient clinically worsened as the steroids were tapered, an examination of his CSF revealed a WBC count of 17 cells/μL. CSF cytology was benign, and flow cytometry showed no evidence of a lymphoproliferative disorder. He was ultimately diagnosed with large B cell lymphoma was ultimately diagnosed by brain biopsy. CSF mNGS detected more than 20 CNVs at a tumor fraction of 22%.

#### Case 138

An adult woman with a history of hypertension, depression, ulcerative colitis, and rheumatoid arthritis on azathioprine presented with 1 year of cognitive decline and gait instability and was found to have multifocal T2 hyperintense bi-frontal, left posterior temporal and parieto-occipital lesions with restricted diffusion on MRI (Figure 3C). An examination of her CSF revealed a WBC count of 18 cells/μL. CSF cytology was benign and flow cytometry was not performed. Intravascular large B cell lymphoma was diagnosed by brain biopsy. CSF mNGS detected one CNV and detected a tumor fraction of 49%.

## Discussion

Transformative advances in NGS and bioinformatics allow us to generate and parse through terabytes of genomic data from patients to find evidence of inherited diseases, cancers, and infections. Because various fields take advantage of similar technological platforms, data generated to make one type of diagnosis (e.g., noninvasive prenatal testing to identify fetal chromosomal abnormalities) can be re-analyzed to detect other disease entities (e.g., maternal cancers^21,22^). Here, we show that data generated by an mNGS assay developed to diagnose infections can be reanalyzed to detect malignant aneuploidy in CSF with moderate sensitivity and high specificity, including in patients with clinical phenotypes that initially raised suspicions for CNS infections.

The detection of solid CNS tumors and CNS lymphomas can be challenging with CSF cytology’s sensitivity ranging from 2-32% in the first CSF sample^23^. In the two presented case-control studies, we detected aneuploidy when cytology and flow cytometry of the CSF were not positive or had ambiguous findings. While the detection of aneuploidy in CSF does not render a specific cancer diagnosis, the high specificity we observed (no false positives in the 84 negative controls) suggests that a positive result could motivate more focused diagnostic testing for a CNS neoplasm in the right clinical context.

Tumor fractions of positive cases were high (median 31-41%) and were occasionally higher than tissue (e.g. Case 44, Figure 2), consistent with previously reported high tumor fractions in CSF^17^. These results suggest that further molecular testing, such as with cancer panels or epigenetic classification^24,25^, has the theortical potential to distinguish between tumor types using CSF. In contrast to cancer somatic mutation panels, the increasingly rapid turnaround time for mNGS, its low DNA input requirements, and the lack of a required enrichment protocol makes it a potentially useful adjunctive tool for cancer detection in CSF that, unlike cytology and flow cytometry, is not dependent on preserving cell integrity. This is especially the case in CNS malignancy patients whose clinical presentations were initially more concerning for an infectious meningitis or meningoencephalitis.

This study has several limitations. These case-control studies were retrospective. In addition, we cannot directly assess the positive and negative predictive value of the assay because the prevalence of CNS malignancy in our study is artificial and not reflective of a relevant clinical population. For the test performance the main source of residual CSF samples mainly through flow cytometry testing introduced a selection bias towards patients who may have lymphoma. Our exclusion of patients with acute leukemia and most cases of systematic cancers limits our ability to assess the false positive rate in patients with coincident systemic malignancy; hence, the test specificity may be lower for those patients. We also did not assess benign tumors or pre-cancers. Thus, this assay’s real-world accuracy needs to be assessed in a larger, prospective cohort study of patients who are referred by their physicians for CSF mNGS with more systematic and long-term follow-up. The clinical utility of the assay could be evaluated if subjects are randomized to have the CNV analysis performed on their mNGS data, especially if no pathogen is identified.

In summary, we believe these preliminary studies, which include provocative individual cases in which tumor detection could have been reached earlier without additional invasive testing, motivate further research in this area.

**Table S1:** Patient and Sample Characteristics in the Test Performance Study and the Suspected Neuroinflammatory Disease Study

| **Test Performance Study** |  |  |
| --- | --- | --- |
| **Patient Demographics** | **Positives (n=55)<sup>a</sup>** | **Negatives (n=70)** |
| Age – years |  |  |
| Median (interquartile range) | 62 (54-71) | 53 (38-66) |
| Range | 27-84 | 0-86 |
| Sex – Female – no. (%) | 23 (41.8%) | 31 (44.3%) |
| History of malignant CNS neoplasm* | 30 (54.5%) | 0 |
| **Sample Characteristics** |  |  |
| Cytology |  |  |
| Positive | 18 (34.5%) | 0 (0.0%) |
| Atypical Cells Present | 10 (18.2%) | 4 (5.7%) |
| Benign | 21 (38.2%) | 37 (52.9%) |
| Not performed/unavailable | 5 (9.1%) | 29 (41.4%) |
| Flow Cytometry |  |  |
| Aberrant clone | 26 (47.3%) | 2 (2.9%) |
| Atypical clone | 4 (7.3%) |  |
| No evidence of lymphoproliferative disorder | 22 (40.0%) | 39 (55.7%) |
| Not performed/unavailable | 3 (5.5%) | 29 (41.4%) |
| Culture Performed | 20 (38.5%) | 47 (69.1%) |
| WBC (cells/uL) |  |  |
| Median (interquartile range) | 5 (1-106.5)<sup>b</sup> | 16 (2-111)<sup>c</sup> |
| Range | 0-495<sup>b</sup> | 0-13,000<sup>c</sup> |
| Imaging |  |  |
| Leptomeningeal | 18 (10.5%) |  |
| Lesion abuts CSF space | 42 (75.0%) |  |
| Unknown | 1 (1.8%) |  |
| **Suspected Neuroinflammatory Study** |  |  |
| **Patient Demographics** | **Positives (n = 12)<sup>a</sup>** | **Negatives (n = 17)** |
| Age (years) |  |  |
| Median (interquartile range) | 71.5 (63.5-74.8) | 44 (33-62) |
| Range | 47-83 | 5-71 |
| Gender |  |  |
| Female – no. (%) | 3 (25%) | 9 (52.9%) |
| Male – no. (%) | 9 (75%) | 8 (47.1%) |
| **Sample Characteristics** |  |  |
| Cytology |  |  |
| Positive | 0 (0.0%) | 0 (0.0%) |
| Atypical Cells Present | 2 (16.7%) | 0 (0.0%) |
| Benign | 9 (75.0%) | 7 (41.2%) |
| Not performed/unavailable | 1 (8.3%) | 17 (58.8%) |
| Flow Cytometry |  |  |
| Aberrant/Atypical clone | 0 (0.0%) | 0 (0.0%) |
| No evidence of lymphoproliferation | 8 (66.7%) | 9 (52.9%) |
| Not performed/unavailable | 4 (33.3%) | 8 (47.1%) |
| Culture Performed (n = 10 data available) | 9 (90.0%) | 7 (41.2%) |
| History of Cancer | 2 (16.7%) | 2 (11.8%) |
| WBC (10<sup>6</sup>/L) |  |  |
| Median (interquartile range) | 12 (4.75-18.0) | 7 (2-14.3)<sup>d</sup> |
| Range | 0-128 | 0-325<sup>d</sup> |
| Imaging |  |  |
| Leptomeningeal | 4 (36.0%) |  |
| Lesion abuts CSF space | 7 (64.0%) |  |
| Unknown | 1 (9.1%) |  |

a – two patients overlapped between the two case-control studies: Case 54/137 and 37/138.
b – WBC from 3 positives were not available
c – WBC from 6 negatives were not available
d – WBC from 3 negatives were not available
\* - History of CNS neoplasm previous to the presentation period from each sample was obtained

## Methods

### Ethics Statement and Sample Collection

CSF samples from the Test Performance case-control study were originally collected at the UCSF Clinical Laboratories between 2017-2019 as part of routine clinical testing and retrospectively enrolled through protocols approved by the UCSF Institutional Review Board (18-25287). For the Suspected Neuroinflammatory Disease study, written and informed consent was obtained from UCSF patients with suspected neuroinflammatory disease (or surrogate representatives) who enrolled in an IRB-approved prospective study between 2014-2019 at UCSF protocols (13-12236).

### Sample selection

#### Test Performance Case-Control Study

In the first study, CSF specimens were sent to the UCSF Clinical Laboratories (San Francisco, California, United States) for diagnostic testing between 2017 and 2019. Positive and negative controls were retrospectively identified through continuous screening for the following inclusion and exclusion criteria.

Positive controls were from patients with either a primary or metastatic CNS malignancy, excluding leukemia. CSF samples were considered “positive” if the sample was positive by either 1) CSF cytology with malignant cells or 2) CSF flow cytometry consistent with a clonal, malignant cell population. Samples with negative cytology and flow cytometry were placed into a separate positive control category if a tumor was eventually confirmed by either 1) positive CSF cytology/cytometry in another sample, 2) a tissue diagnosis, or 3) Neuroradiology and Oncology diagnostic consensus with intention to treat. Samples were excluded if they came from patients already undergoing active chemotherapy or radiation therapy unless cytology and flow cytometry of the CSF was positive.

The negative controls were CSF specimens from patients who ultimately received a clinical diagnosis of a CNS infection or autoimmune disease and no known malignant cancer diagnosis unless the patient had paraneoplastic disease or had been in remission for > 10 years. Cases without a definitive diagnosis upon follow-up were excluded as they did not fit into positive or negative categories.

Most specimens (80% of the 105 samples with records) were continuously received from flow cytometry testing, and the rest were from hematology and microbiology screened based on positive results from cytology or microbiological testing based on a previous study enrollment^12^. The number of positive and negative controls ultimately chosen was a convenience sample based on the maximum number of samples available, not based on a pre-specified power calculation.

#### Suspected Neuroinflammatory Disease Case-Control Study

In the second study, adult cancer cases and controls were identified from a prospective study of CSF mNGS for suspected neuroinflammatory disease. These patients were referred between 2014 to 2019. Cases were identified by retrospective chart review.

Cases for whom a definitive diagnosis was never obtained or who had a benign tumor were excluded. Patients with an eventual diagnosis of an autoimmune disorder were included as negative controls. Cases that were clinically determined (and confirmed with long term follow-up) to be of solely paraneoplastic causes were not excluded.

### Neuroimaging Analysis

To be able to correlate CSF cancer detection with the MRI brain findings of the subjects in the CSF Test Performance and Suspected Neuroinflammatory Disease studies, all available MRIs were reviewed by a board-certified neuroradiologist (A.R.) who was blinded to the mNGS results and recorded the number of lesions, location of the lesions, presence or absence of leptomeningeal involvement, and whether at least one lesion abutted the CSF space.

### Cerebrospinal Fluid Sample Extraction

For the first case-control study (Test Performance), residual CSF specimens were collected from the UCSF Clinical Laboratories (Hematology and Flow cytometry, Chemistry, and Microbiology). The retrospective archival material was kept at 4°C for up to 2 weeks and then processed as described below before freezing. For the second study (Suspected Neuroinflammatory Disease), CSF specimens were collected directly from patients or residual CSF was collected from the clinical laboratory. Samples were either immediately extracted or frozen at −80°C as the original samples or in storage buffer (20% glycerol, 20 mM HEPES, 0.02% NaN3 in PBS) at a 1:1 sample:buffer volumetric ratio.

Original or thawed CSF was centrifuged at 3000*g* −16,000*g* for 10 minutes and the supernatant was stored at −80 degrees Celsius. Total nucleic acid extraction was performed by the EZ1 Advanced XL BioRobot using the EZ1 Virus Mini Kit v2.0 (QIAGEN) with 400µL CSF input and 60µL extracted DNA output.

### Sequencing Library Preparation

Whole genome sequencing (WGS) library preparation was performed using the NEBNext Ultra II DNA Library Prep Kit (New England Biolabs) on an epMotion 5075 liquid handler (Eppendorf) using the manufacturer’s protocol unless otherwise stated. All reagent volumes were halved, and the input was also halved to 25 µL of extracted DNA.

For bead purification, we used Ampure XP beads (Beckman Coulter). PCR amplification of the adapter ligated DNA was up to 26 cycles using the manufacturer’s protocol and, the cDNA libraries were dual indexed before pooling. Sequencing was performed on an Illumina HiSeq 1500/2500 or Nextseq 550 in either the single-end or optionally paired-end for 140 bp.

### Tumor Tissue Extraction and Library Preparation

Formalin fixed paraffin blocks were used to obtain correlated CNV data from cancer tissue obtained from the same subject. All archival tissue was no longer needed for clinical care. A neuropathologist (T.T.) identified regions of high tumor content on correlated tissue section(s). A disposable dermal punch was used to either punch out or scrape tissue from regions of interest. DNA was extracted from the fixed tissue using the Quick-DNA FFPE Miniprep kit (Zymo Research). Each sample was sheared using focused acoustics to approximately 250 bp in a microTUBE (Covaris) and quantified on a spectrometer (Nanodrop, Thermo Fisher). Up to 50 ng of sheared DNA was used for WGS library preparation as described above.

### Bioinformatics

Raw data was demultiplexed to raw FASTQ files and adapter trimmed with cutadapt (v1.16). The mNGS analysis pipeline used SURPI^8,26^ to make microbial identifications. The copy number variations were called by aligning to the human genome hg38 and de-duplicating reads with BWA^27^ (v0.7.12). CNVkit^28^ (v0.9.1) was used to segment the genome into bins, display a log2 copy number across all bins, and calculate a median line (orange in plots). Sequencing data from CSF samples were normalized to a reference from an asymptomatic, healthy male plasma control. Sequencing data derived from the tumor blocks were normalized to a resected tonsil tissue from an otherwise healthy male with an oropharyngeal infection. All CNV calls and calculated tumor fractions were made based on blinded interpretation of the log2 copy ratio plots by a board-certified molecular pathologist (W.G.). In keeping with the interpretation of aneuploidy from copy ratio plots clinically and from past experience with plasma and other body fluids (data submitted but not published), only large (>10 Mbp) CNVs or whole chromosome arms larger than that size elevated or depressed at an even level of copy ratio were called. Frequent, low-level, and small deviations from the baseline of log2 copy ratio of 0 were interpreted as an artifact based on noise. Sex chromosomes and areas near telomeres and centromeres known for routine copy number changes were not used. Chromosome 19, known for high GC content and high degrees of noise^29,30^, was not used to call CNVs, but was used as a metric for the degree of background noise.

### Statistical Analysis

After the the mNGS assay was assessed for its ability to identify CNV in CSF, and these results were compared to the clinicial and laboratory evidence for each patient, test performance characteristics, including sensitivity and specificity, were calculated along with 95% confidence intervals. Sensitivity and specificity confidence intervals were calculated using the Clopper-Pearson exact method.

### Data Availability

Raw sequencing data is available on SRA under BioProject PRJNA678570 for all patients who consented to genomic data sharing. All other metadata, including copy ratio plots, raw data linked to those plots, and human-subtracted metagenomics data are located at: https://doi.org/10.5281/zenodo.4682169.

## Data Availability

Data Availability
Raw sequencing data is available on SRA under BioProject PRJNA678570 for all patients who consented to genomic data sharing. All other metadata, including copy ratio plots, raw data linked to those plots, and human-subtracted metagenomics data are located at: https://doi.org/10.5281/zenodo.4682169.

https://www.ncbi.nlm.nih.gov/bioproject/?term=678570

## Author Contributions

Drs Gu and Wilson had full access to all of the data in the study and take responsibility for the integrity of the data and the accuracy of the data analysis.

Study concept and design: Gu, Wilson

Patient Recruitment: Zorn, Sample, Cree, Douglas, Richie, Shah, Josephson, Gelfand, Gold, Wang.

Sample acquisition and sequencing preparation: Gu, Hsu, Sucu, Zorn, Arevalo, Gopez, Gottschall, Wang, Tihan.

Acquisition, analysis, or interpretation of data: Gu, Rauschecker, Hsu, Zorn, Arevalo, Gopez, Federman, Talevich, Nguyen.

Drafting of the manuscript: Gu, Rauschecker, Wilson

Critical revision of the manuscript for important intellectual content: All authors. Statistical analysis: Gu.

Administrative, technical, or material support: Gu, Miller, Wang, Tihan, DeRisi, Chiu, Wilson

Study supervision: Gu, Wang, DeRisi, Wilson.

## Acknowledgements

We thank the patients in this study for donating critical CSF samples to research. We thank the UCSF clinical laboratory staff for assistance with sample collection, in particular members of the Clinical Immunology, Microbiology, and Hematology laboratories.

## References

1. Glaser CA, Honarmand S, Anderson LJ, et al. Beyond Viruses: Clinical Profiles and Etiologies Associated with Encephalitis. Clin Infect Dis. 2006;43(12):1565–1577. doi:10.1086/509330

2. Granerod J, Tam CC, Crowcroft NS, Davies NWS, Borchert M, Thomas SL. Challenge of the unknown. A systematic review of acute encephalitis in non-outbreak situations. Neurology. 2010;75(10):924–932. doi:10.1212/WNL.0b013e3181f11d65

3. Granerod J, Ambrose HE, Davies NW, et al. Causes of encephalitis and differences in their clinical presentations in England: a multicentre, population-based prospective study. Lancet Infect Dis. 2010;10(12):835–844. doi:10.1016/S1473-3099(10)70222-X

4. Wilson MR, Naccache SN, Samayoa E, et al. Actionable Diagnosis of Neuroleptospirosis by Next-Generation Sequencing. New England Journal of Medicine. 2014;370(25):2408–2417. doi:10.1056/NEJMoa1401268

5. Wilson MR, O’Donovan BD, Gelfand JM, et al. Chronic Meningitis Investigated via Metagenomic Next-Generation Sequencing. JAMA Neurol. Published online April 16, 2018. doi:10.1001/jamaneurol.2018.0463

6. Wilson MR, Shanbhag NM, Reid MJ, et al. Diagnosing Balamuthia mandrillaris Encephalitis With Metagenomic Deep Sequencing. Annals of neurology. 2015;78(5):722–730.

7. Wilson MR, Sample HA, Zorn KC, et al. Clinical Metagenomic Sequencing for Diagnosis of Meningitis and Encephalitis. New England Journal of Medicine. 2019;380(24):2327–2340. doi:10.1056/NEJMoa1803396

8. Miller S, Naccache SN, Samayoa E, et al. Laboratory validation of a clinical metagenomic sequencing assay for pathogen detection in cerebrospinal fluid. Genome Res. Published online April 16, 2019. doi:10.1101/gr.238170.118

9. Simner PJ, Miller HB, Breitwieser FP, et al. Development and Optimization of Metagenomic Next-Generation Sequencing Methods for Cerebrospinal Fluid Diagnostics. J Clin Microbiol. 2018;56(9). doi:10.1128/JCM.00472-18

10. Wang S, Chen Y, Wang D, et al. The Feasibility of Metagenomic Next-Generation Sequencing to Identify Pathogens Causing Tuberculous Meningitis in Cerebrospinal Fluid. Front Microbiol. 2019;10:1993. doi:10.3389/fmicb.2019.01993

11. Piantadosi A, Kanjilal S, Ganesh V, et al. Rapid Detection of Powassan Virus in a Patient With Encephalitis by Metagenomic Sequencing. Clin Infect Dis. 2018;66(5):789–792. doi:10.1093/cid/cix792

12. Gu W, Deng X, Lee M, et al. Rapid pathogen detection by metagenomic next-generation sequencing of infected body fluids. Nat Med. 2021;27(1):115–124. doi:10.1038/s41591-020-1105-z

13. Pan W, Gu W, Nagpal S, Gephart MH, Quake SR. Brain Tumor Mutations Detected in Cerebral Spinal Fluid. Clinical Chemistry. 2015;61(3):514–522. doi:10.1373/clinchem.2014.235457

14. Wang Y, Springer S, Zhang M, et al. Detection of tumor-derived DNA in cerebrospinal fluid of patients with primary tumors of the brain and spinal cord. Proc Natl Acad Sci U S A. 2015;112(31):9704–9709. doi:10.1073/pnas.1511694112

15. De Mattos-Arruda L, Mayor R, Ng CKY, et al. Cerebrospinal fluid-derived circulating tumour DNA better represents the genomic alterations of brain tumours than plasma. Nature Communications. 2015;6:8839. doi:10.1038/ncomms9839

16. Pentsova EI, Shah RH, Tang J, et al. Evaluating Cancer of the Central Nervous System Through Next-Generation Sequencing of Cerebrospinal Fluid. J Clin Oncol. 2016;34(20):2404–2415. doi:10.1200/JCO.2016.66.6487

17. Seoane J, Mattos-Arruda LD, Rhun EL, Bardelli A, Weller M. Cerebrospinal fluid cell-free tumour DNA as a liquid biopsy for primary brain tumours and central nervous system metastases. Annals of Oncology. 2019;30(2):211–218. doi:10.1093/annonc/mdy544

18. Taylor AM, Shih J, Ha G, et al. Genomic and Functional Approaches to Understanding Cancer Aneuploidy. Cancer Cell. 2018;33(4):676-689.e3. doi:10.1016/j.ccell.2018.03.007

19. Talevich E, Shain AH, Botton T, Bastian BC. CNVkit: Genome-Wide Copy Number Detection and Visualization from Targeted DNA Sequencing. PLOS Computational Biology. 2016;12(4):e1004873. doi:10.1371/journal.pcbi.1004873

20. Fan HC, Blumenfeld YJ, Chitkara U, Hudgins L, Quake SR. Noninvasive diagnosis of fetal aneuploidy by shotgun sequencing DNA from maternal blood. PNAS. 2008;105(42):16266–16271. doi:10.1073/pnas.0808319105

21. Dharajiya NG, Grosu DS, Farkas DH, et al. Incidental Detection of Maternal Neoplasia in Noninvasive Prenatal Testing. Clinical Chemistry. Published online January 1, 2017:clinchem.2017.277517. doi:10.1373/clinchem.2017.277517

22. Ji X, Li J, Huang Y, et al. Identifying occult maternal malignancies from 1.93 million pregnant women undergoing noninvasive prenatal screening tests. Genetics in Medicine. Published online April 12, 2019:1. doi:10.1038/s41436-019-0510-5

23. Scott BJ, Douglas VC, Tihan T, Rubenstein JL, Josephson SA. A Systematic Approach to the Diagnosis of Suspected Central Nervous System Lymphoma. JAMA Neurol. 2013;70(3):311–319. doi:10.1001/jamaneurol.2013.606

24. Miller AM, Shah RH, Pentsova EI, et al. Tracking tumour evolution in glioma through liquid biopsies of cerebrospinal fluid. Nature. 2019;565(7741):654–658. doi:10.1038/s41586-019-0882-3

25. Capper D, Jones DTW, Sill M, et al. DNA methylation-based classification of central nervous system tumours. Nature. 2018;555(7697):469–474. doi:10.1038/nature26000

26. Naccache SN, Federman S, Veeeraraghavan N, et al. A cloud-compatible bioinformatics pipeline for ultrarapid pathogen identification from next-generation sequencing of clinical samples. Genome Res. Published online June 4, 2014. doi:10.1101/gr.171934.113

27. Li H, Durbin R. Fast and accurate short read alignment with Burrows-Wheeler transform. Bioinformatics. 2009;25(14):1754–1760. doi:10.1093/bioinformatics/btp324

28. Talevich E, Shain AH, Botton T, Bastian BC. CNVkit: Genome-Wide Copy Number Detection and Visualization from Targeted DNA Sequencing. PLOS Computational Biology. 2016;12(4):e1004873. doi:10.1371/journal.pcbi.1004873

29. Grimwood J, Gordon LA, Olsen A, et al. The DNA sequence and biology of human chromosome 19. Nature. 2004;428(6982):529–535. doi:10.1038/nature02399

30. Fan HC, Blumenfeld YJ, Chitkara U, Hudgins L, Quake SR. Noninvasive diagnosis of fetal aneuploidy by shotgun sequencing DNA from maternal blood. PNAS. 2008;105(42):16266–16271. doi:10.1073/pnas.0808319105

