## Supplemental Materials for "Detection of Neoplasms by Metagenomic Sequencing of Cerebrospinal Fluid"

**Supplementary Materials - Table of Contents**

**Outline:**

**Table S1: Study 1 samples**

**Correlated body fluid and cancer tissue copy ratio plots**

**Table S1: Study 1 samples**

Supplementary Table S1 is located in the file Supplementary_Table_S1.xls

**Correlated body fluid and cancer tissue copy ratio plots**

Sample 1

| CSF | 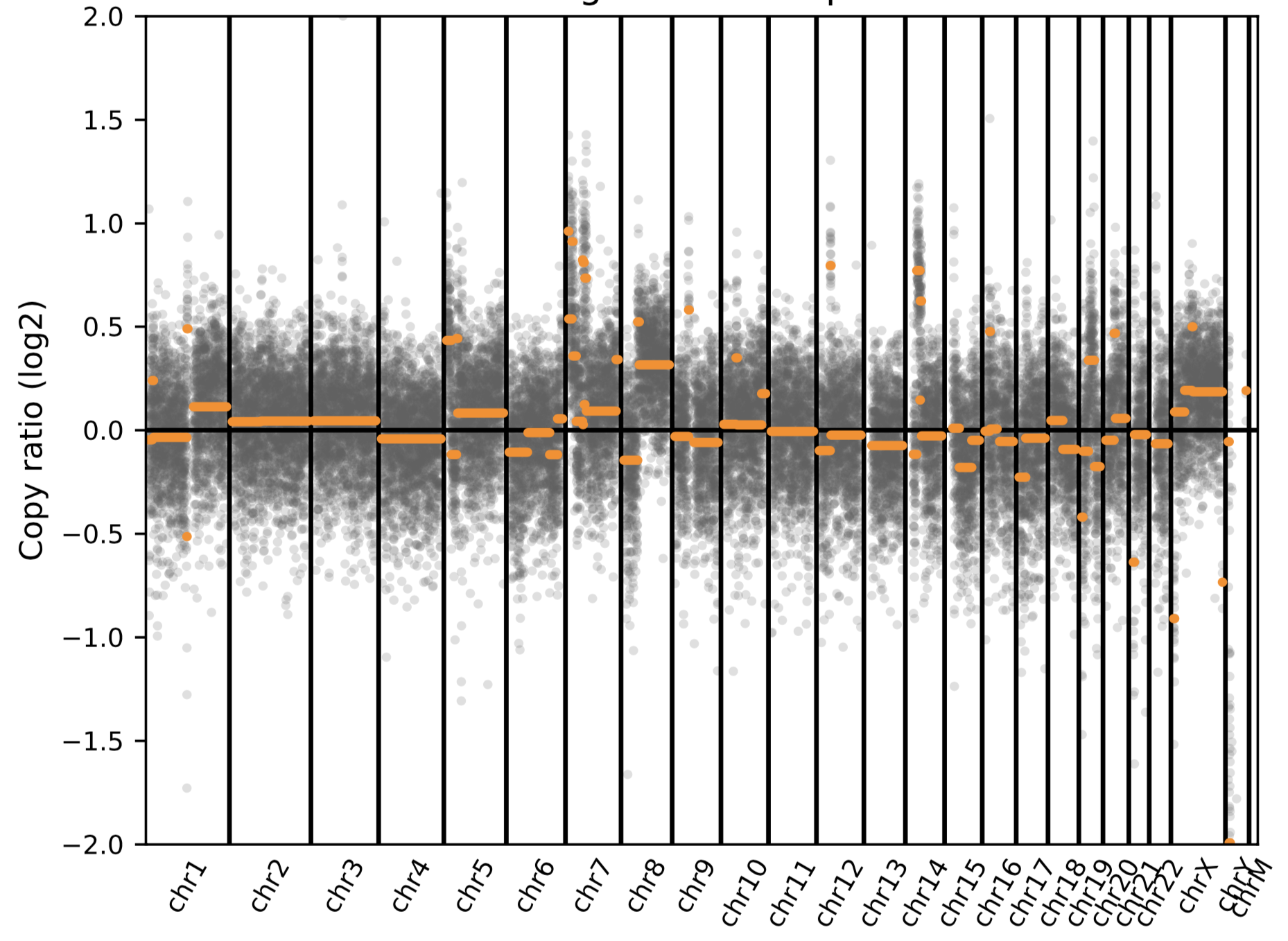 |
| --- | --- |
| Cytology Cell Pellet  (Lung adenocarcinoma) | 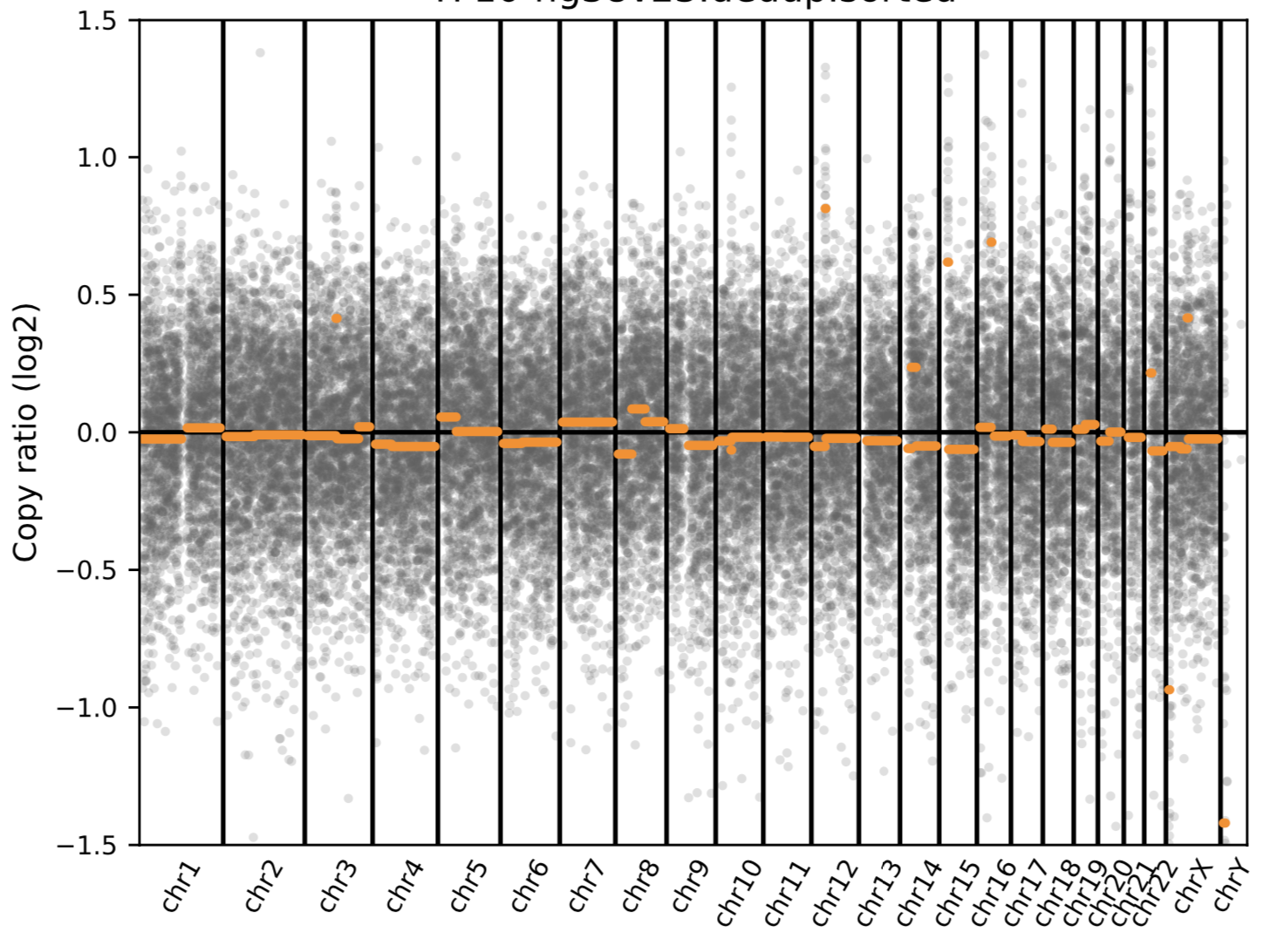 |

Sample 21

| CSF | 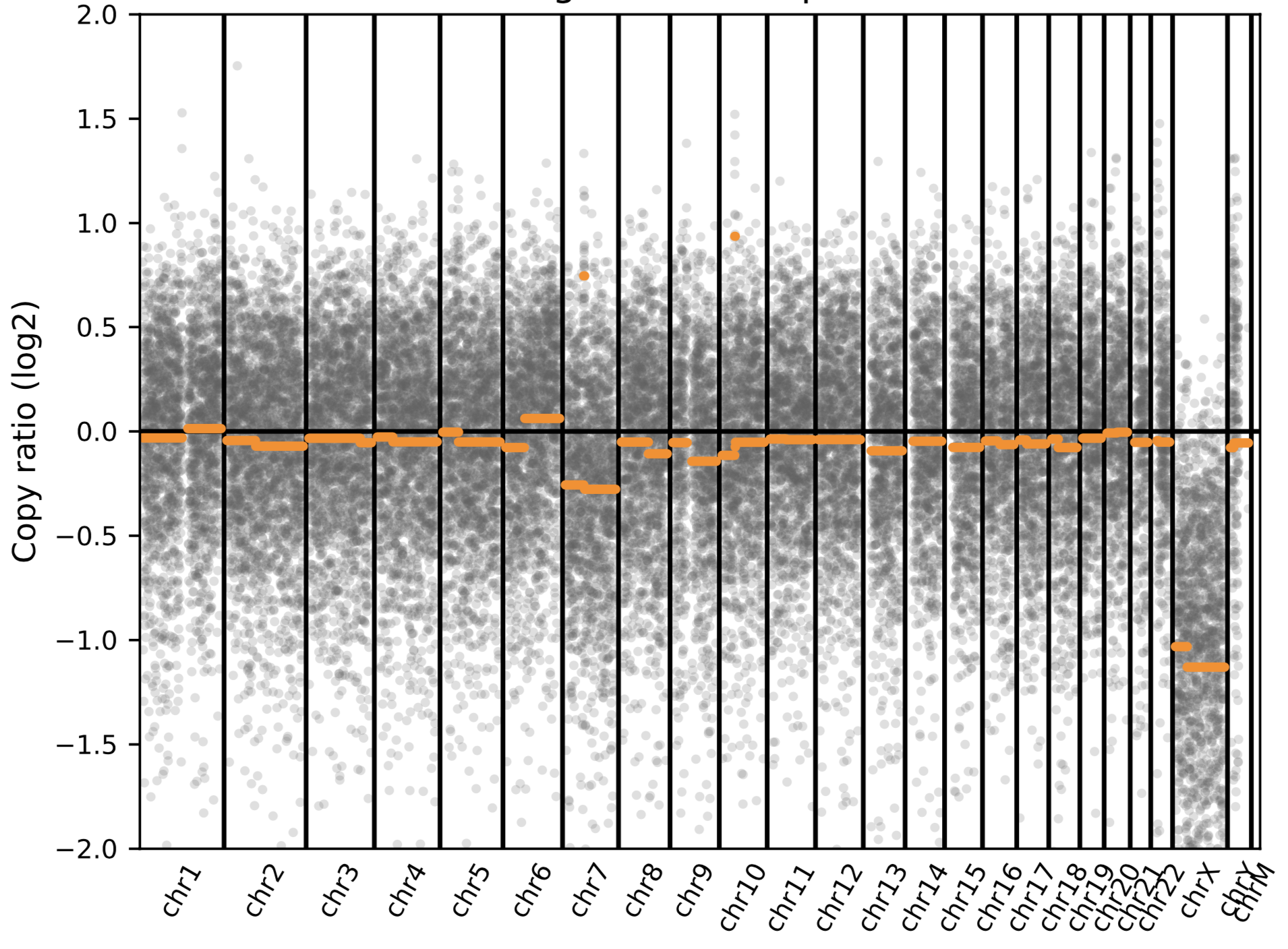 |
| --- | --- |
| Cytogenetics from Bone Marrow  (Chronic Myelogenous Leukemia) | 43,XY,-20,-21,-22[1]  44,XY,-5,-7[1] |

Sample 32

| CSF | 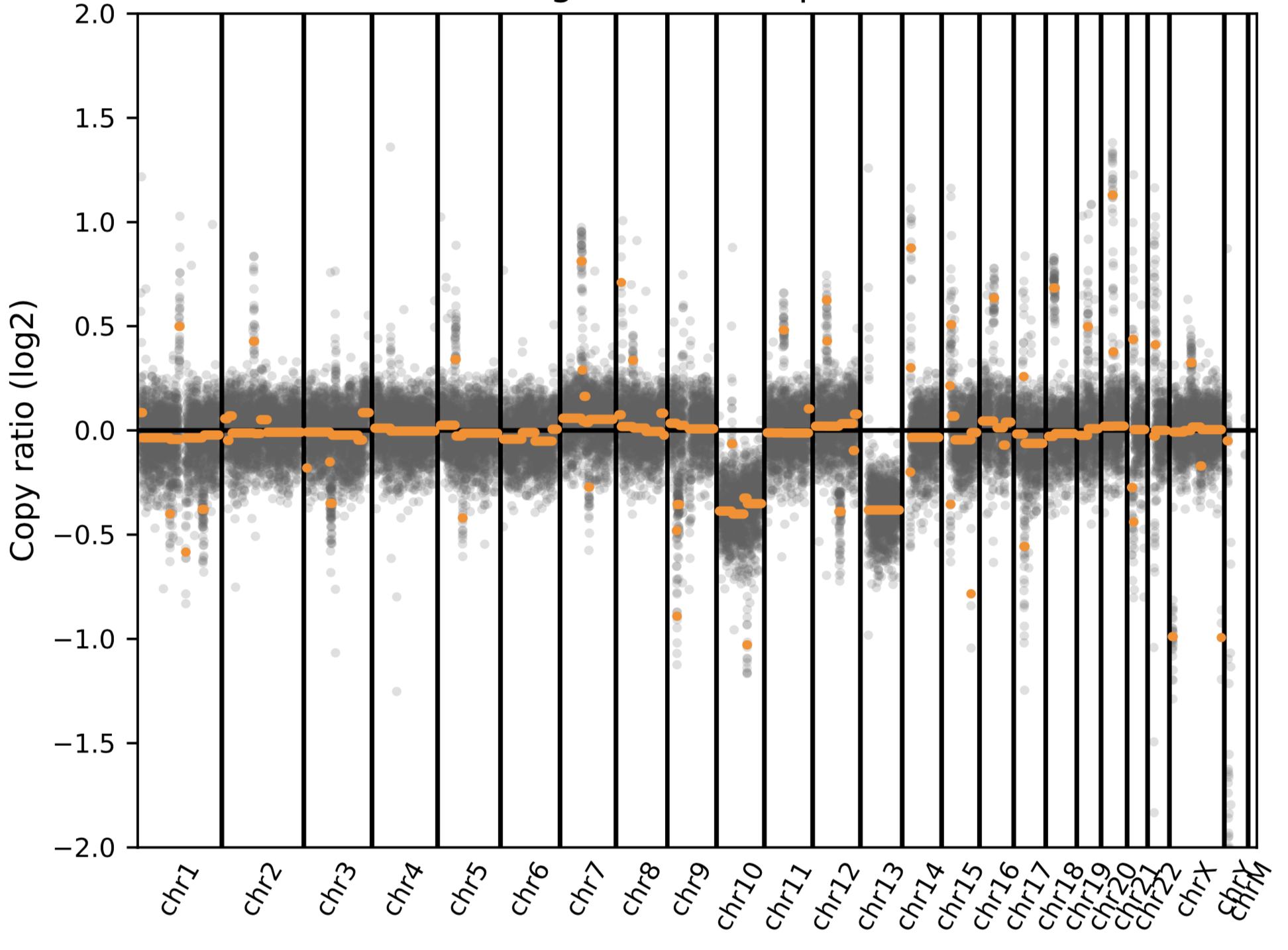 |
| --- | --- |
| Cancer tissue from  Brain biopsy  (Glioblastoma, grade IV) | 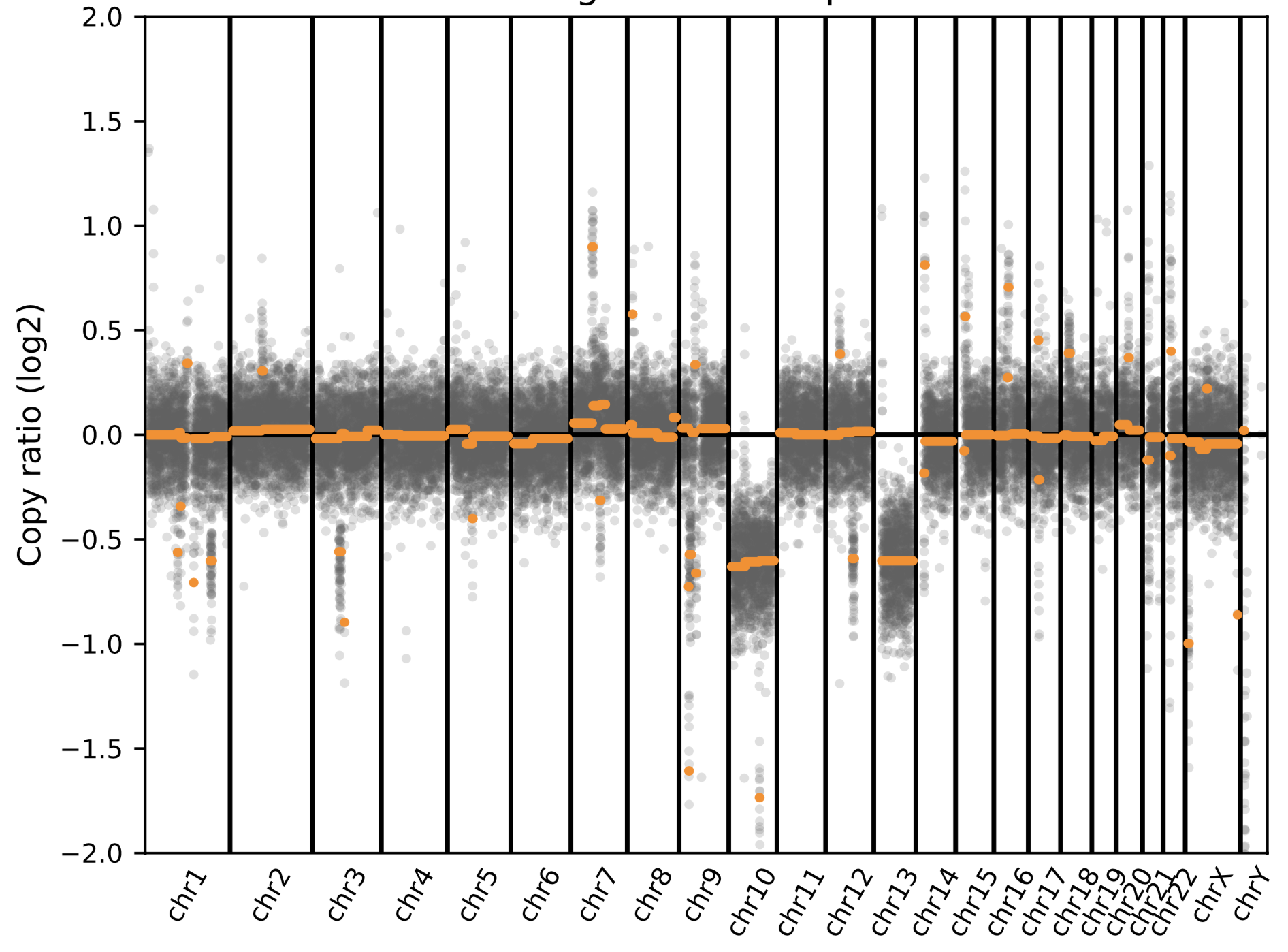 |

Sample 34

| CSF | 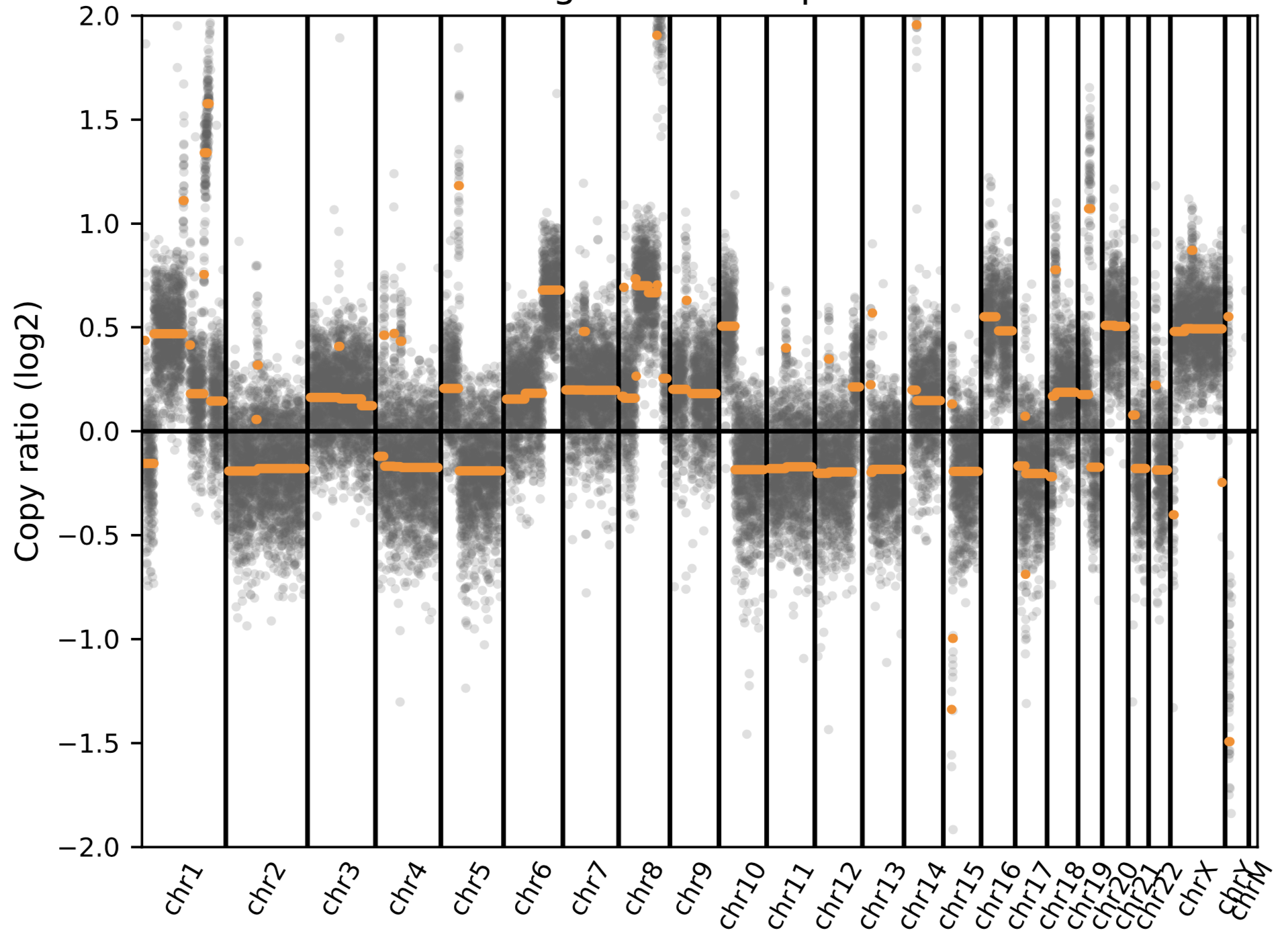 |
| --- | --- |
| Cancer tissue from mastectomy  (Metastatic breast cancer) | 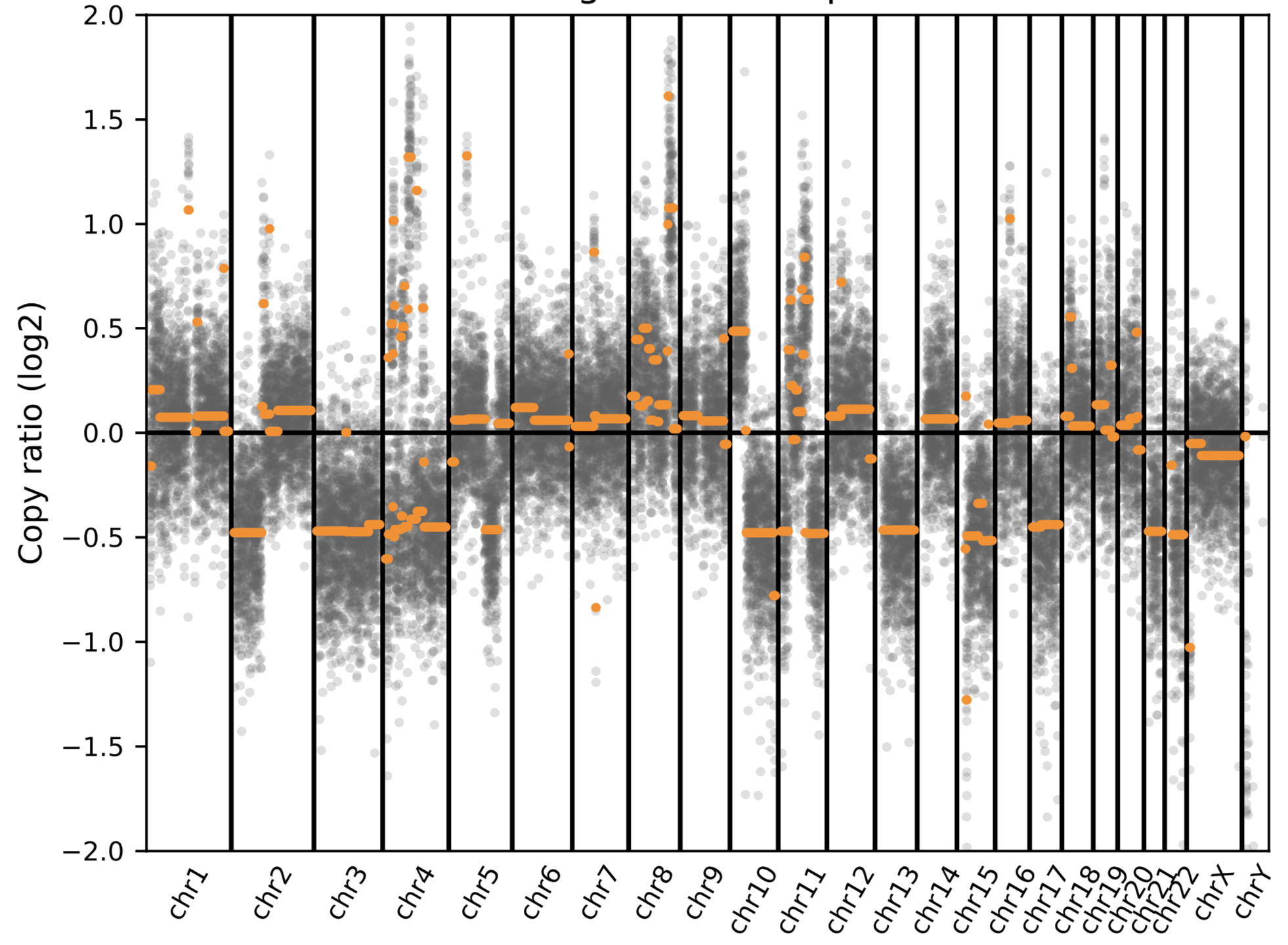 |

Sample 36

| CSF | 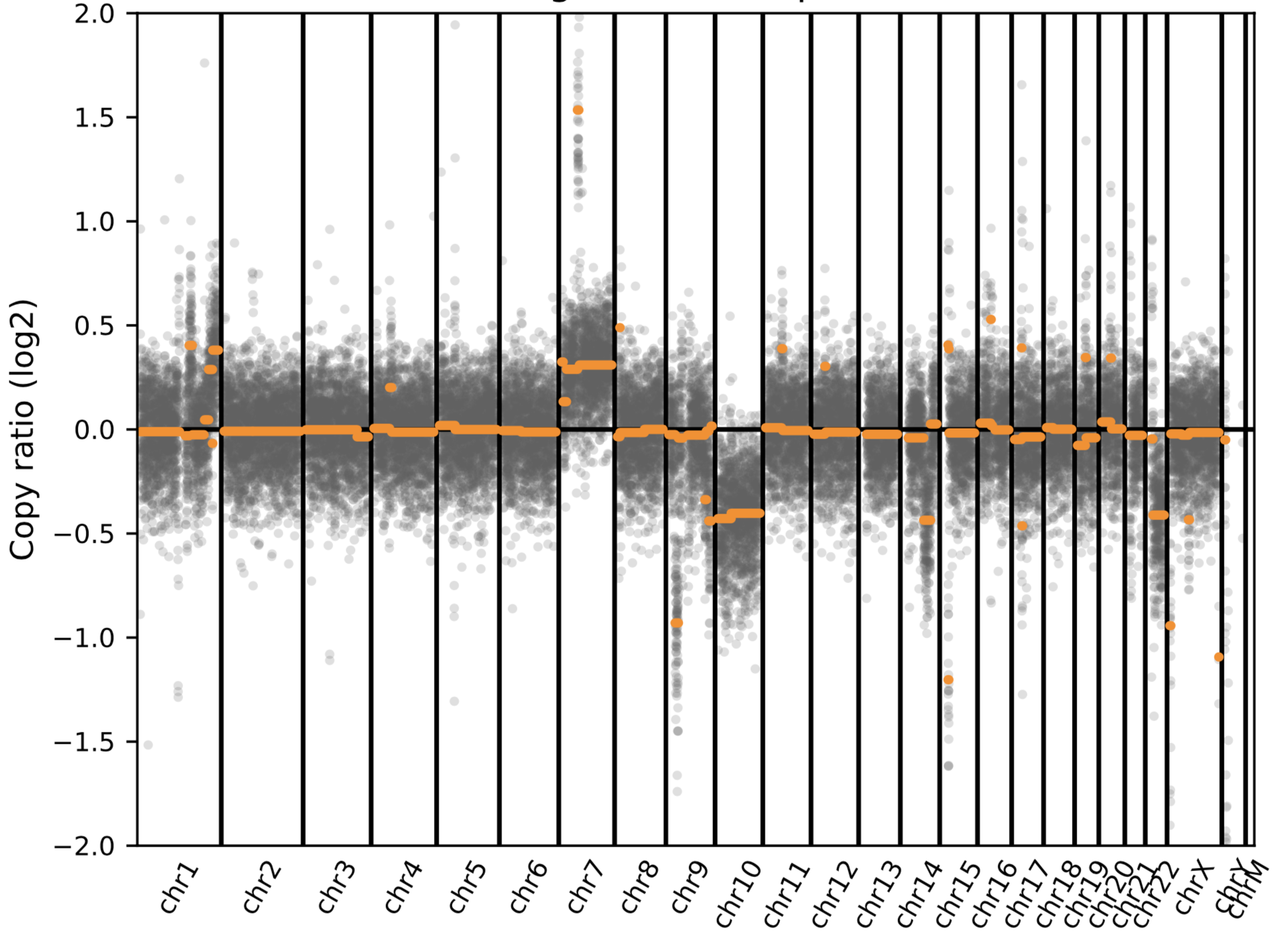 |
| --- | --- |
| Cancer tissue from  Brain biopsy  (Glioblastoma, grade IV) | 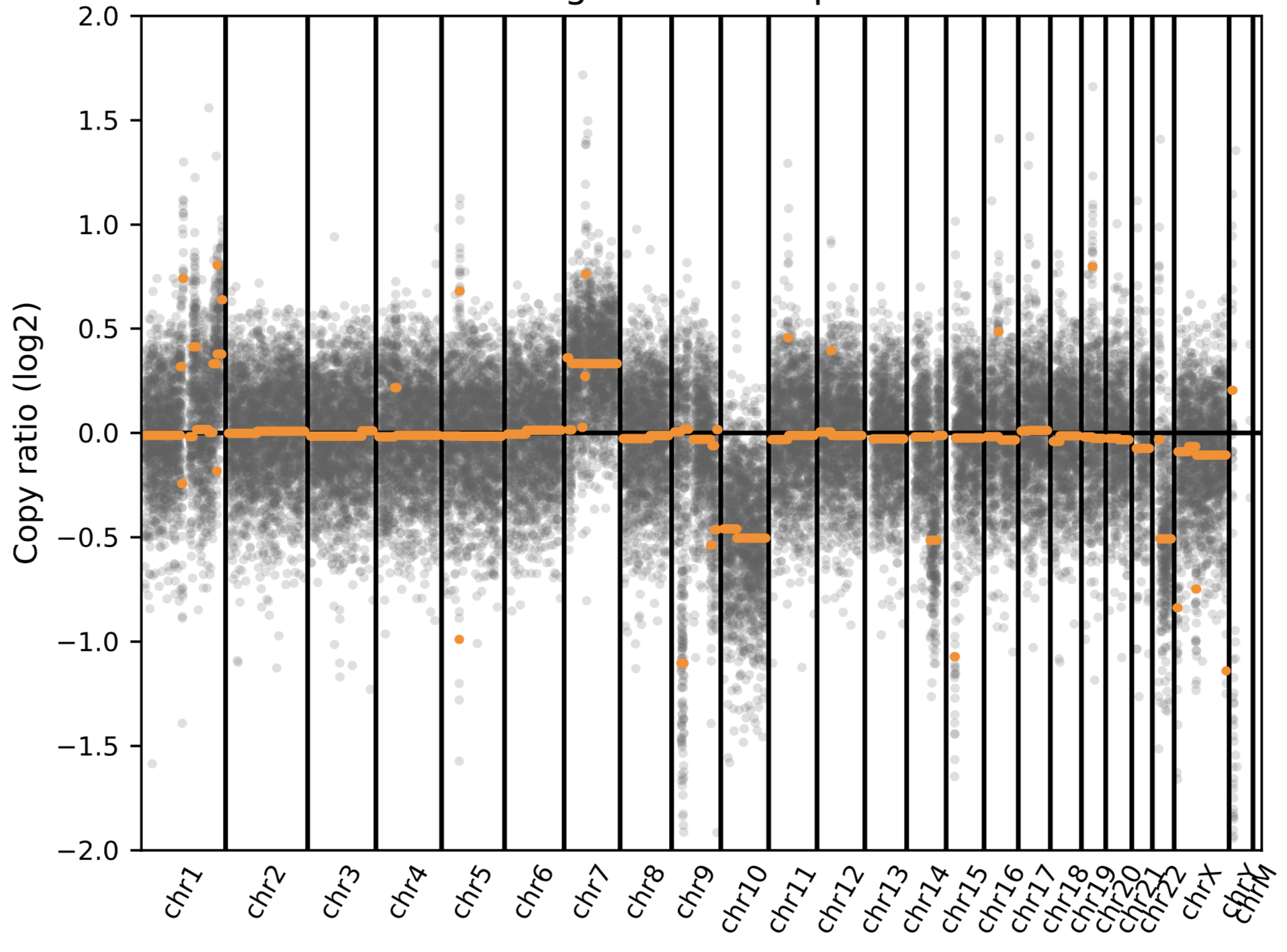 |

Sample 40

| CSF  (negative call on NGS) | 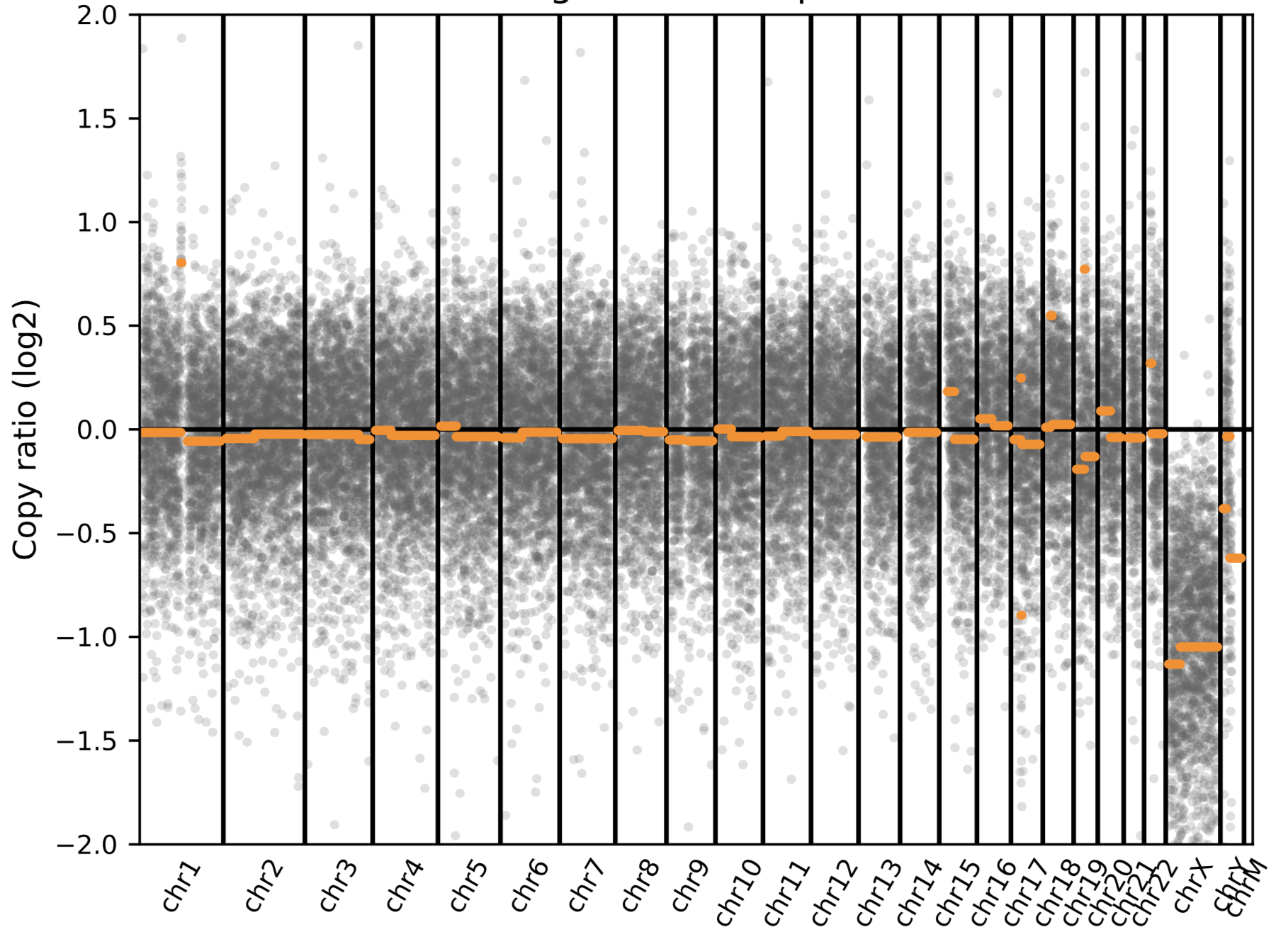 |
| --- | --- |
| Cancer tissue from brain resection | UCSF 500 (cancer panel): focal high level amplification of the MDM4 oncogene on chromosome 1q32. Chromosomal copy number analysis demonstrates gains of 7 and 19, as well as losses of interstitial 8q, distal 9p, 10, 15q, and 18p) |

Sample 8

| CSF | 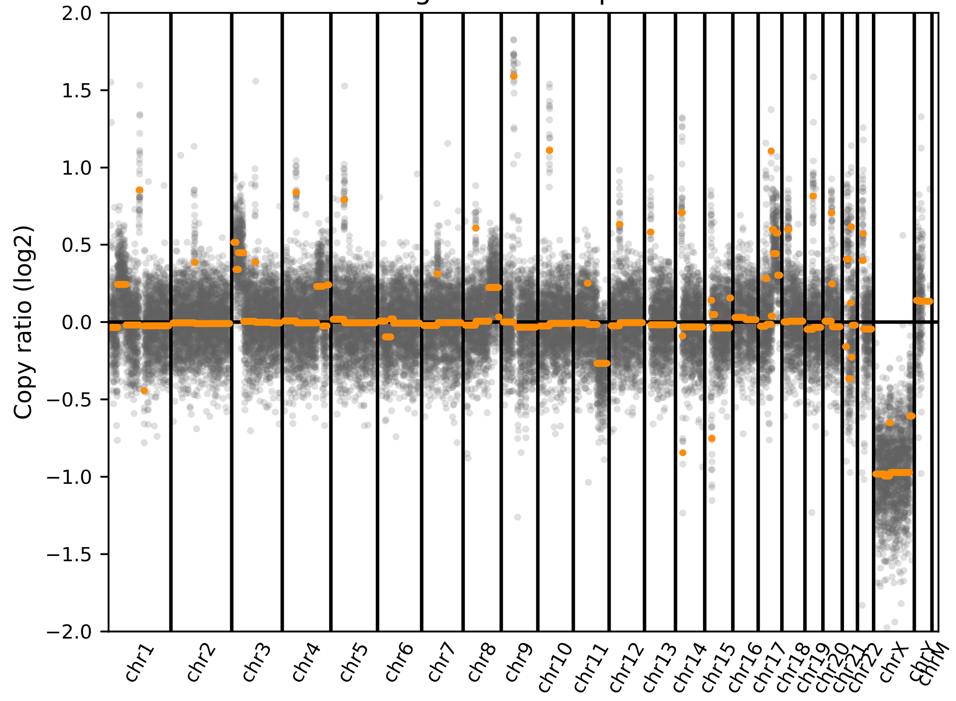 |
| --- | --- |
| Cancer tissue  (T cell lymphoma) | UCSF 500 (cancer panel): gains of chromosome 1p35.2-p31.1, 3p26.3-p22.1, 4q31.1-q32.3, 4q35.1-q35.2, 8q22.2-q24.3, 17q, 21q11.2-q21.1, 21q21.3. Copy loss for 9q (9q21.13, 9q22.32-q22.33, 9q31.2, 9q34.2-q34.3), 11q21-q25, and 21q21.1-  q21.2, 21q22.11. |

Sample 45

| CSF  (Negative call on NGS. Given the tissue correlation data, this false negative was likely due to low tumor fraction in the CSF) | 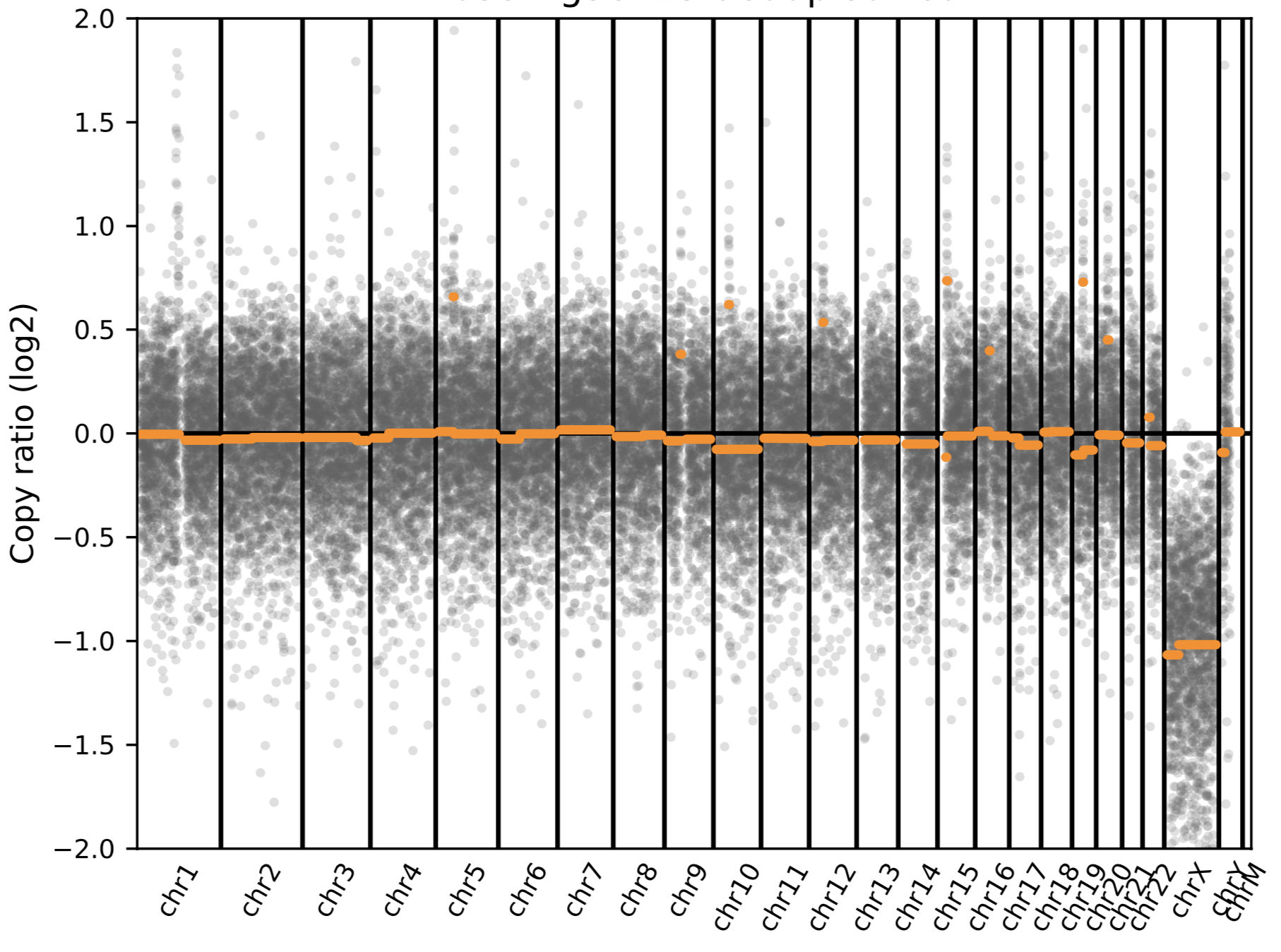 |
| --- | --- |
| Cancer tissue from  Brain biopsy  (Glioblastoma, grade IV) | 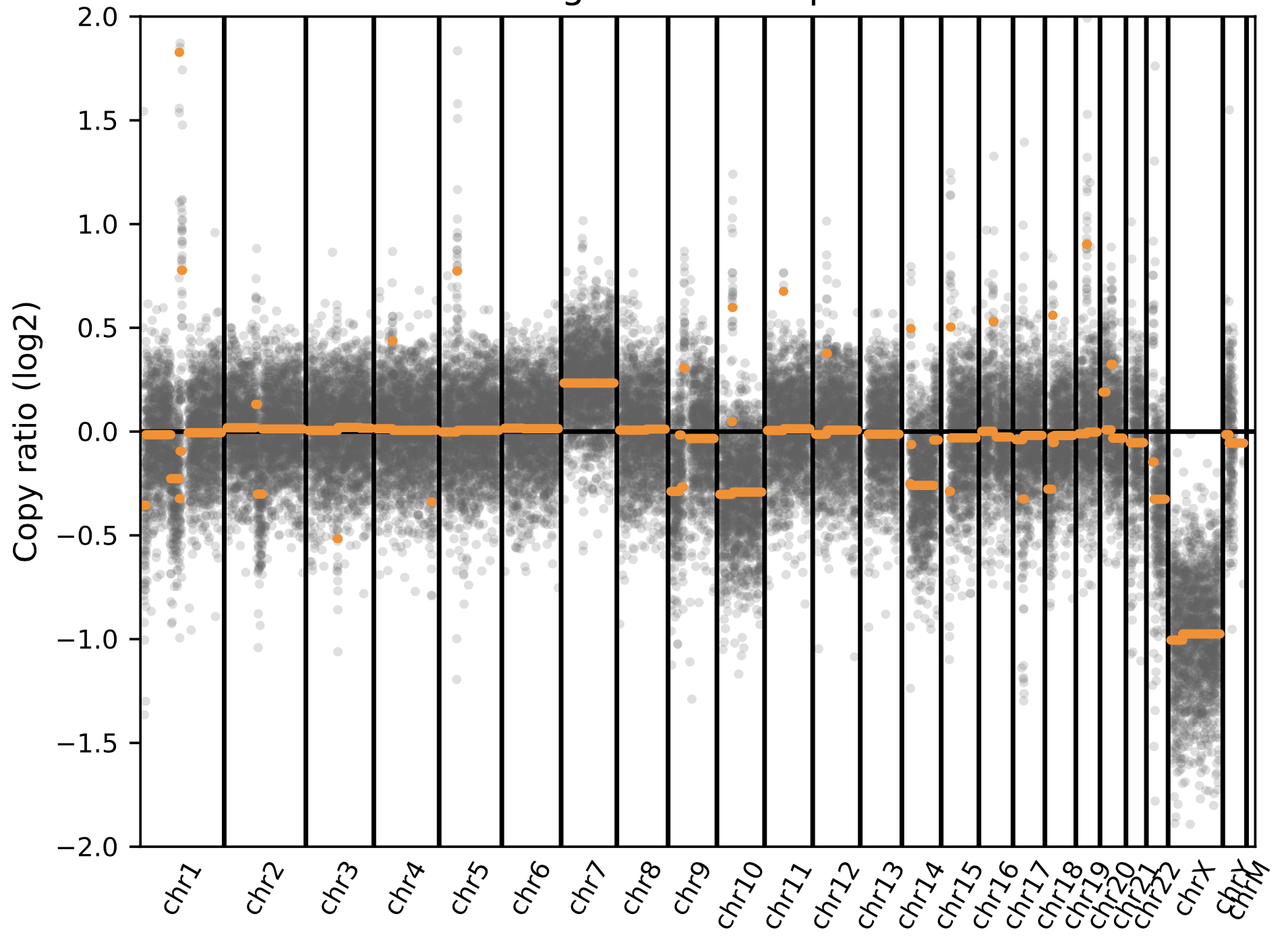 |

Sample 46

| CSF  (Negative call on NGS. Given the tissue correlation data, this false negative was likely due to low tumor fraction in the CSF) | 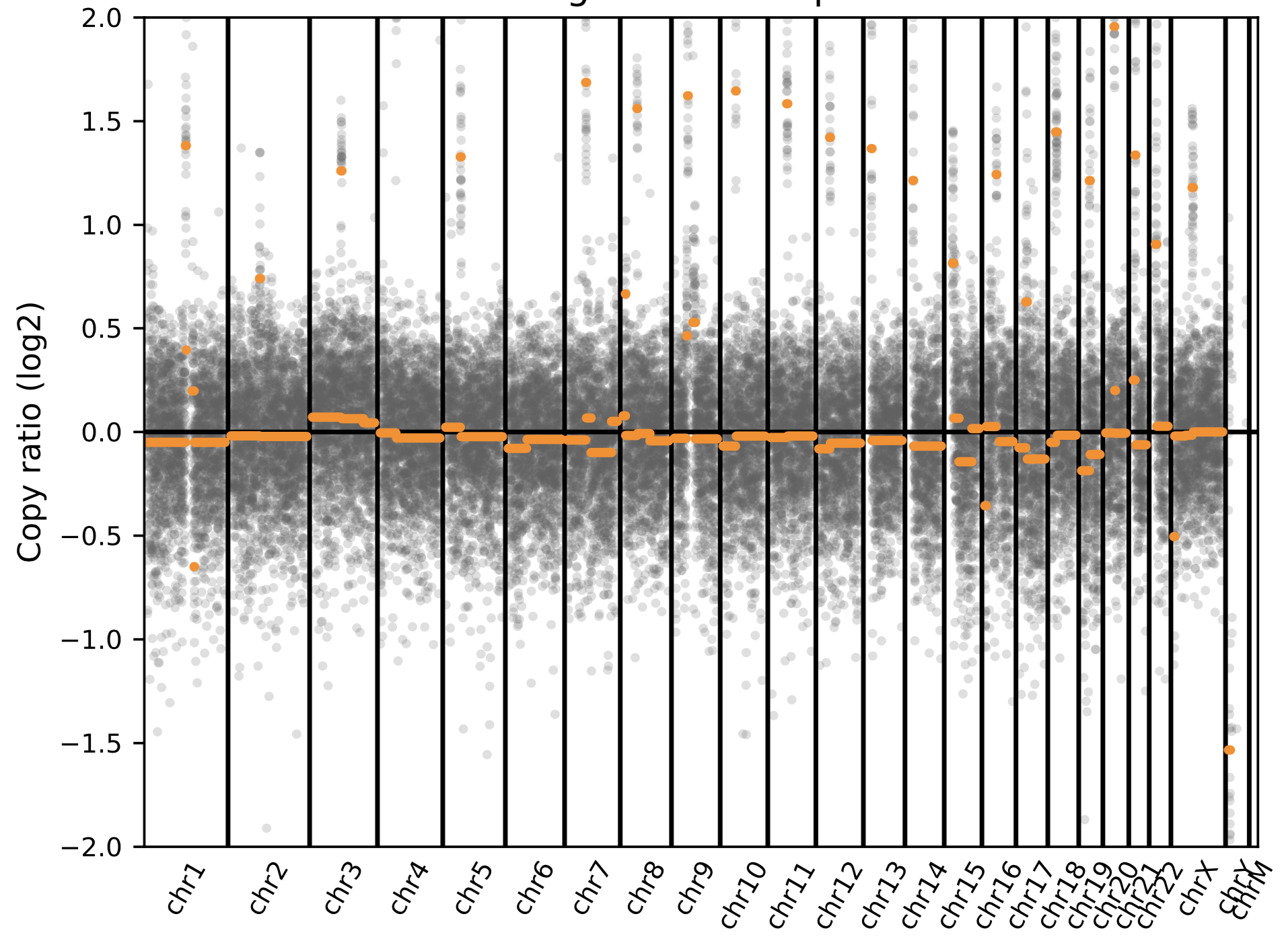 |
| --- | --- |
| Cancer tissue  (low grade marginal zone lymphoma) | 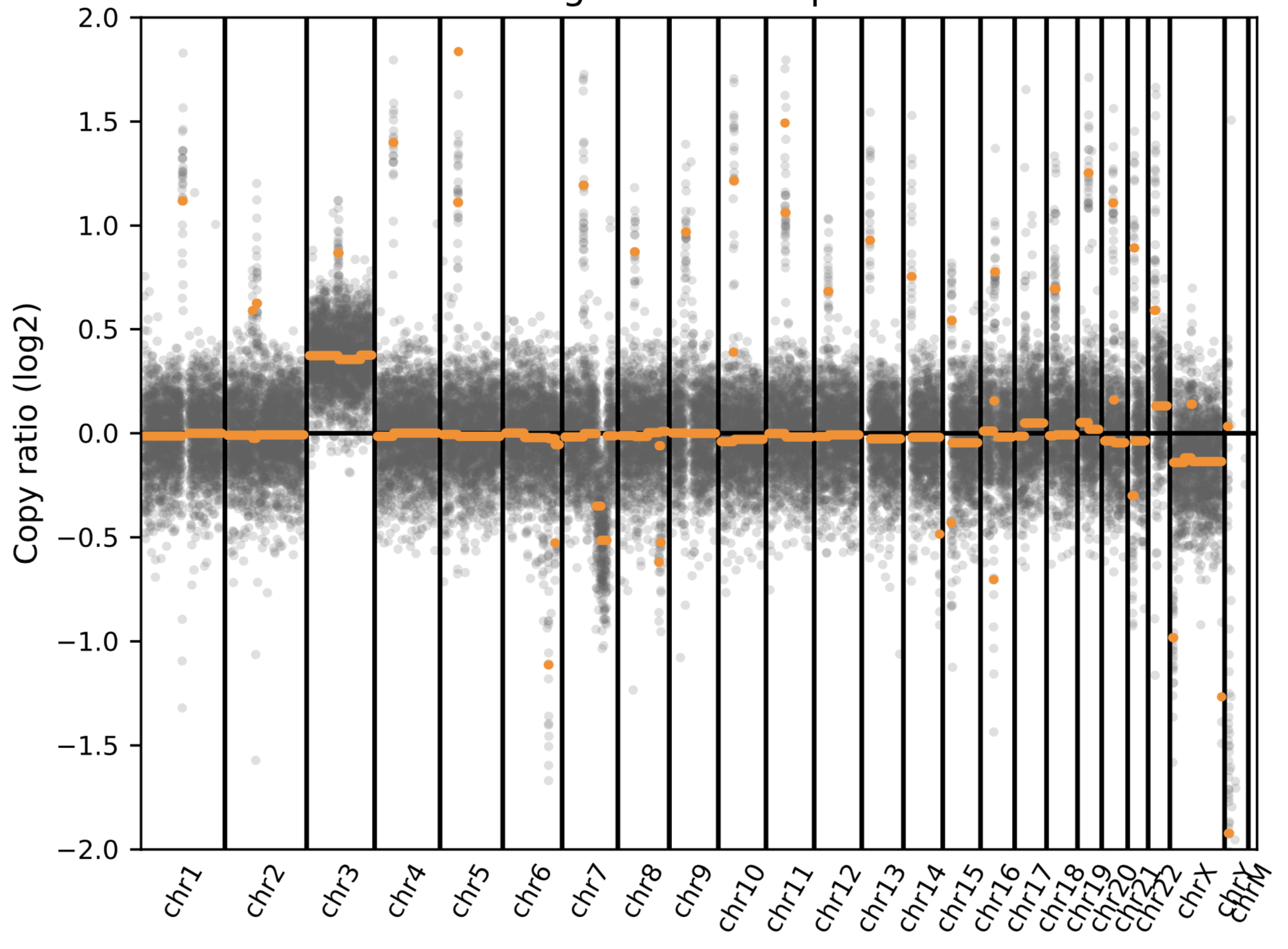 |

Sample 48

| CSF | 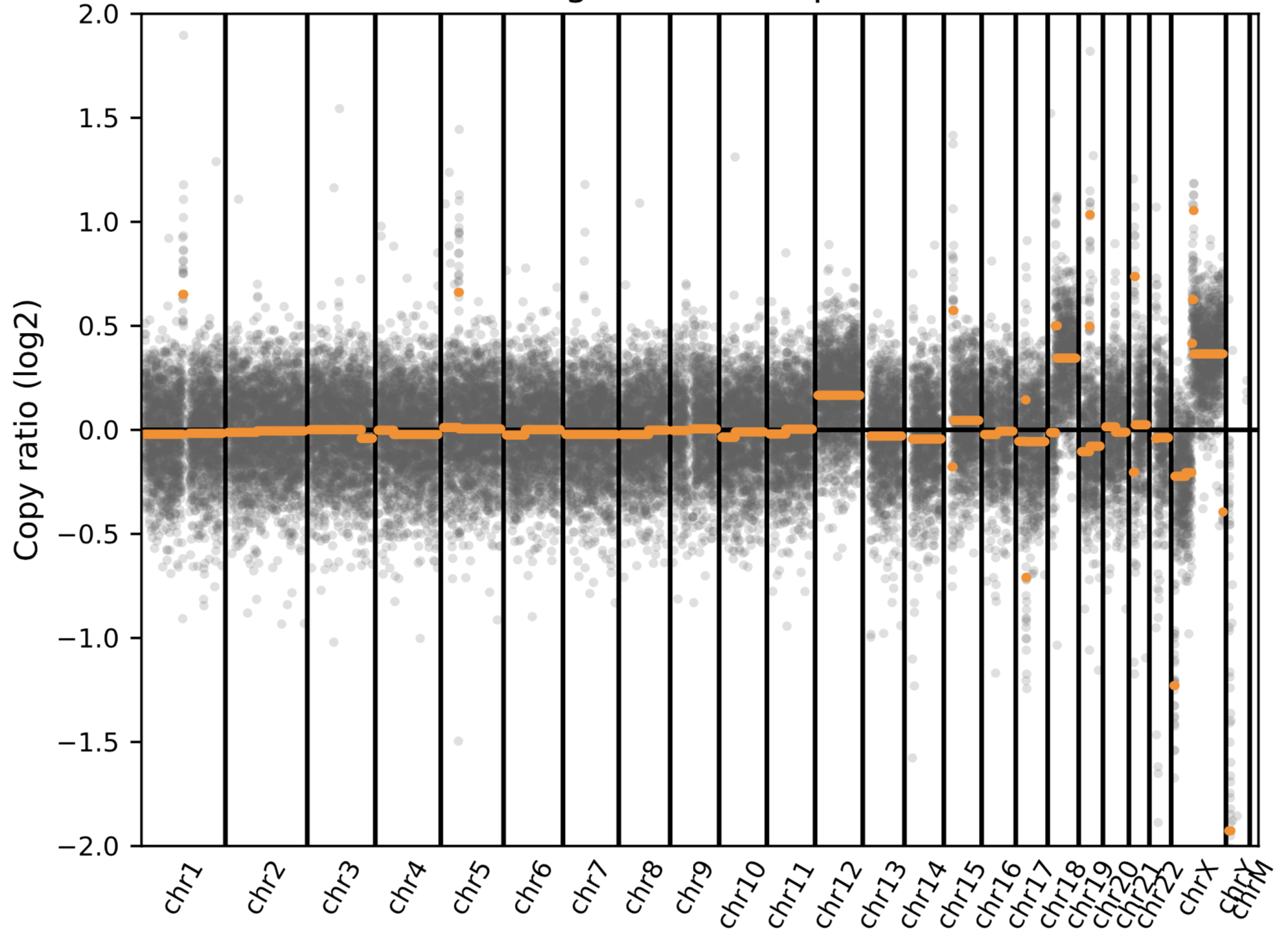 |
| --- | --- |
| Cancer tissue  (large B-cell lymphoma) | 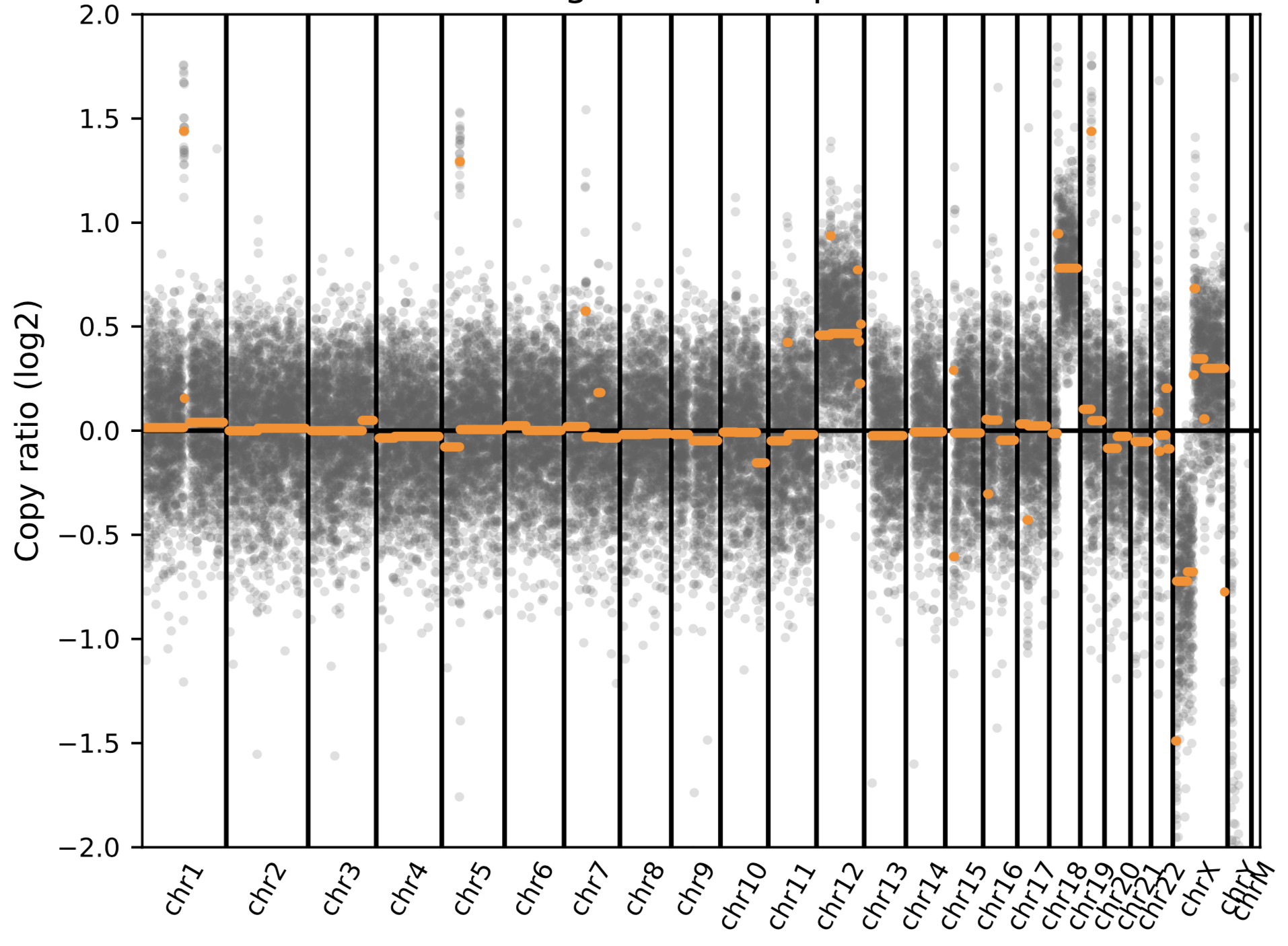 |

Sample 129

| CSF | 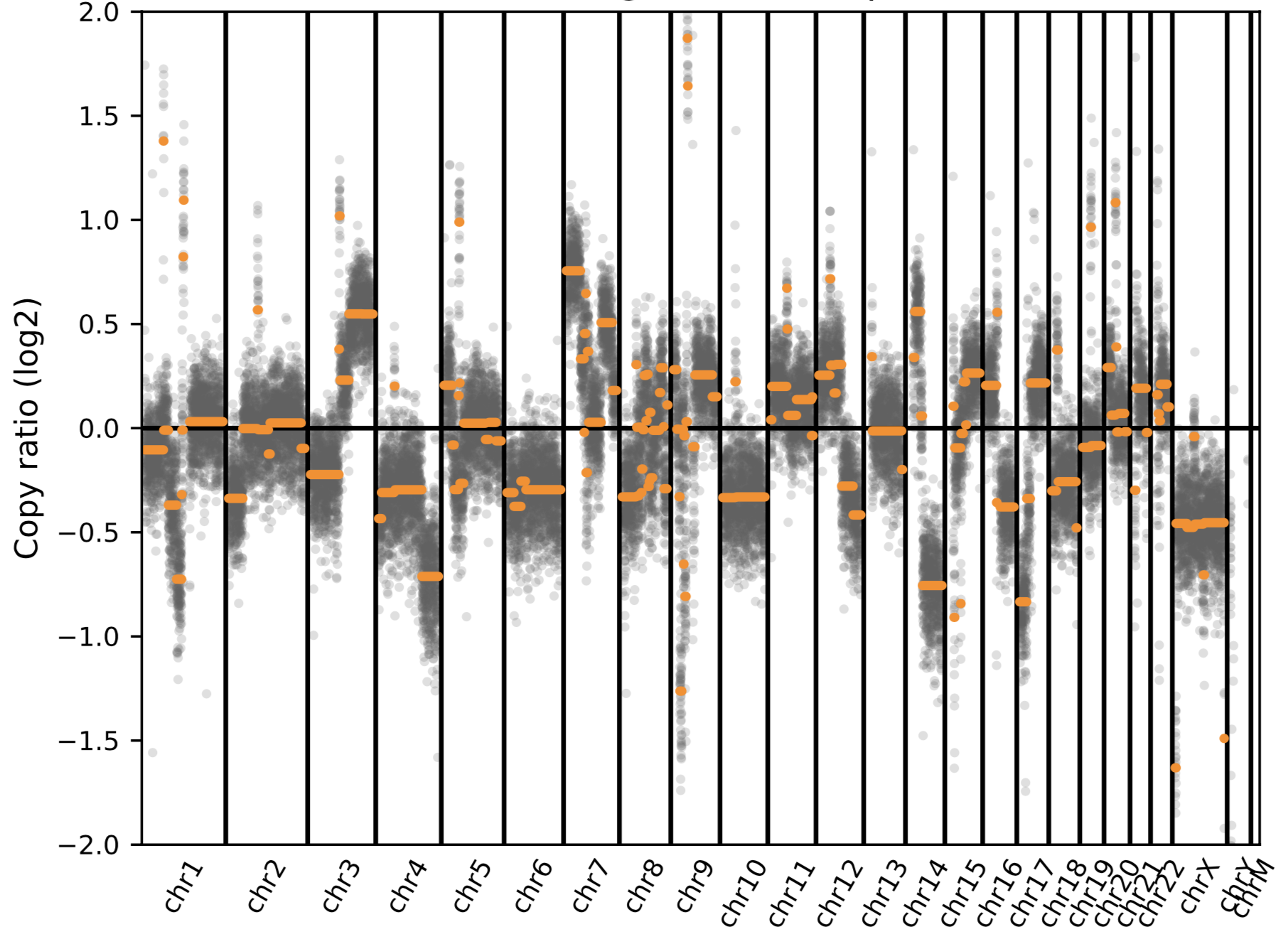 |
| --- | --- |
| Cancer tissue from brain biopsy  (melanoma) | 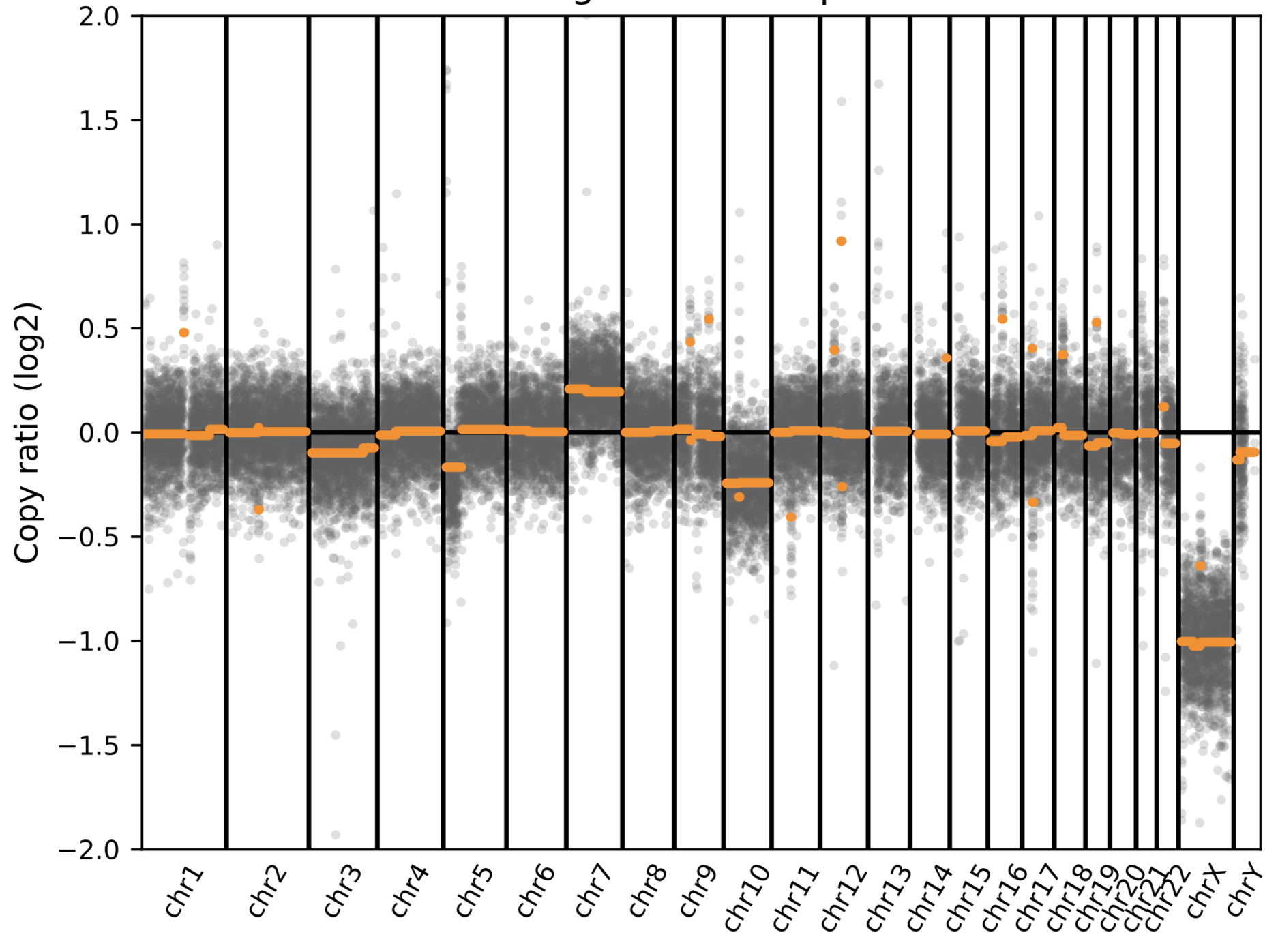 |

Sample 130

| CSF  (negative by mNGS) | 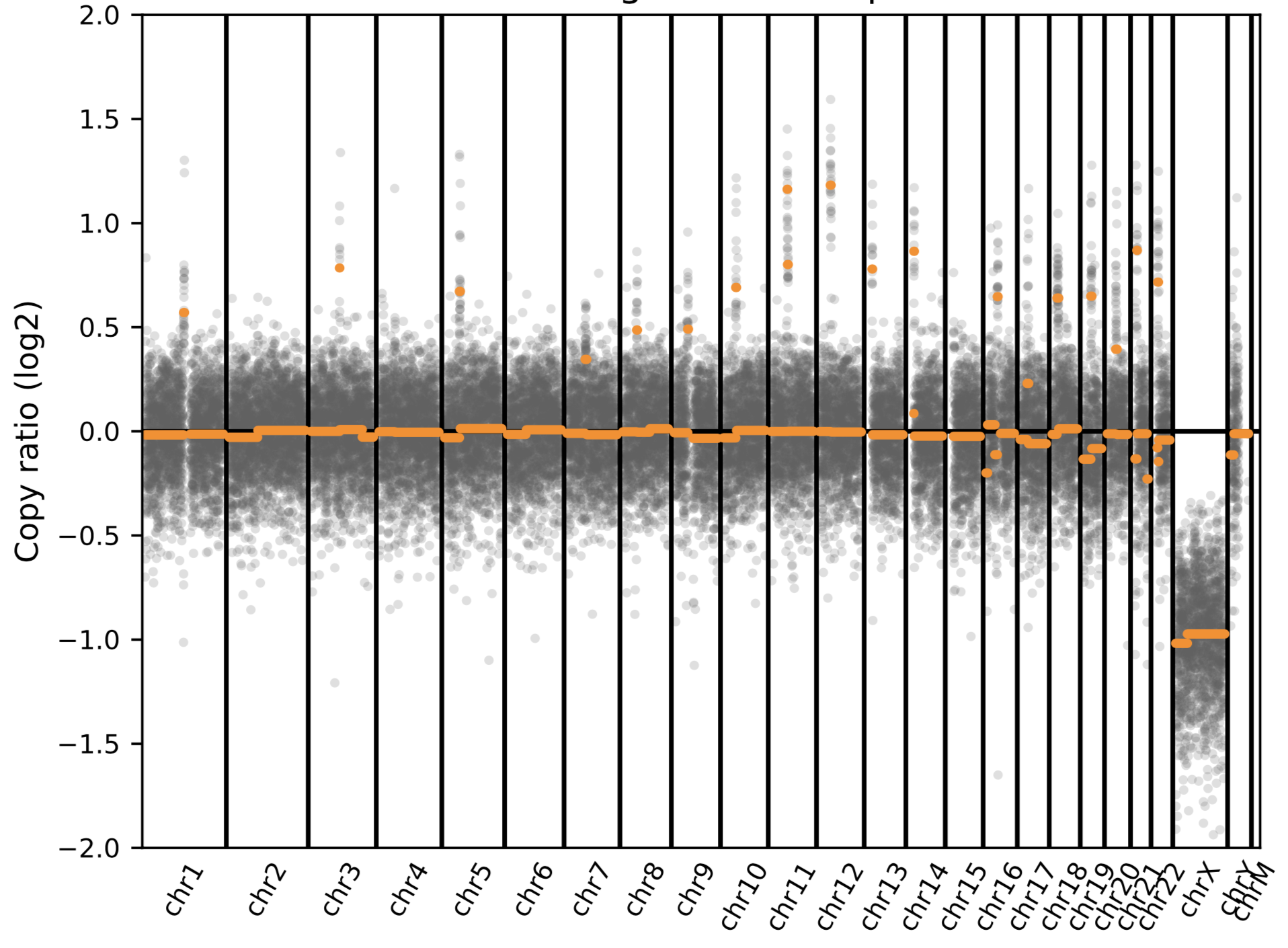 |
| --- | --- |
| Cancer tissue from brain biopsy  (glioblastoma) | 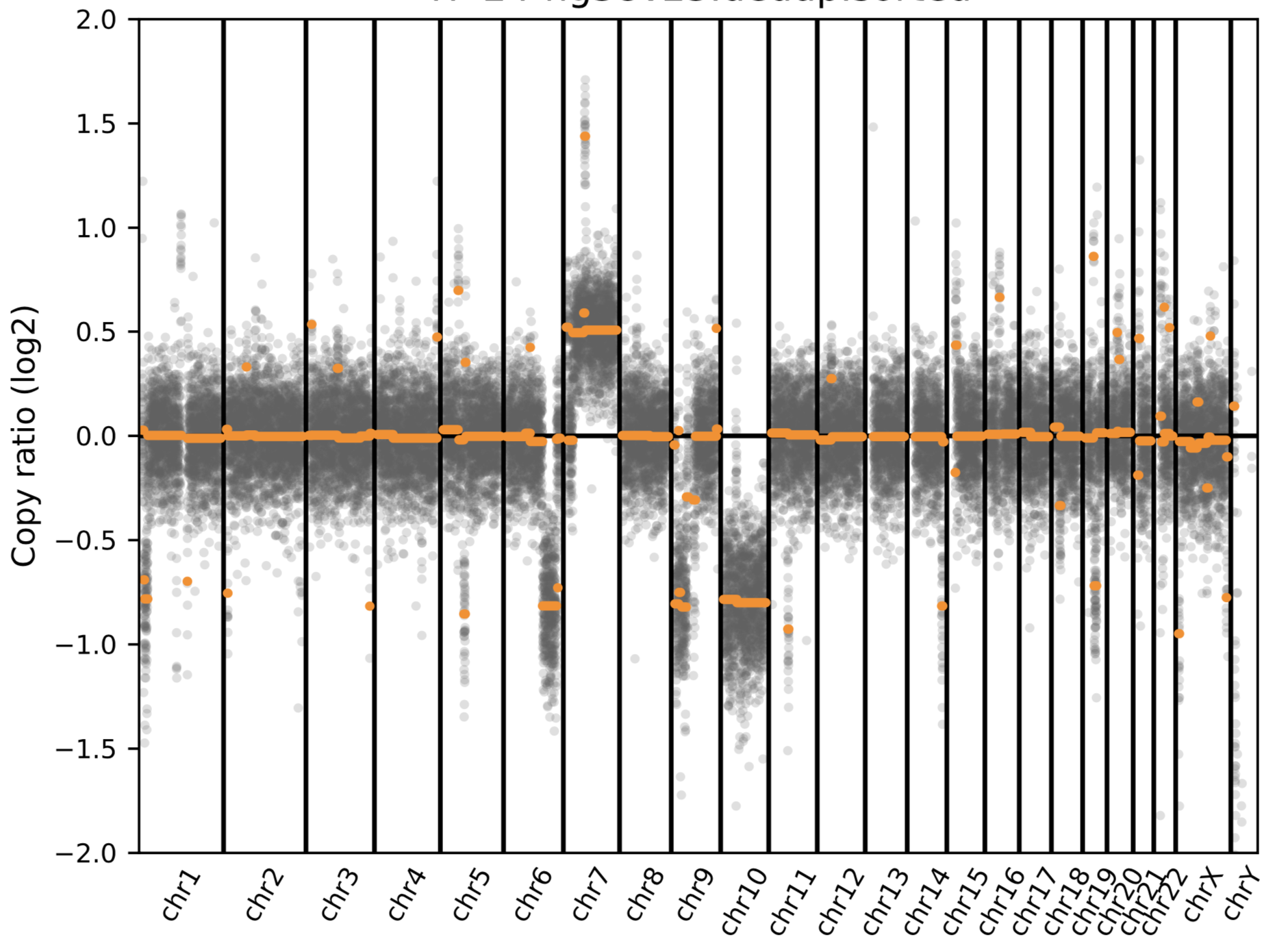 |

Sample 134

| CSF | 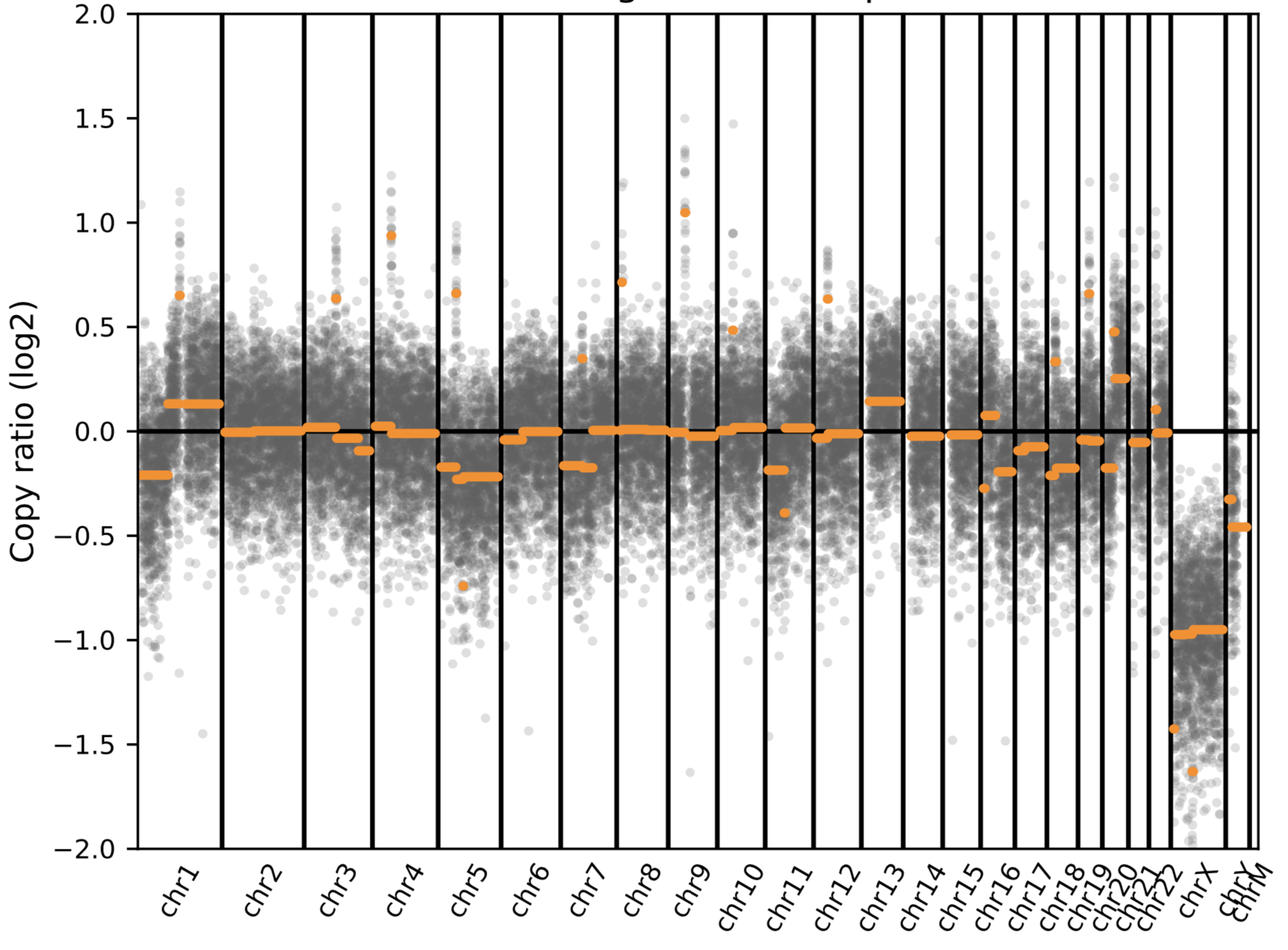 |
| --- | --- |
| Cancer tissue from brain biopsy  (melanoma) | 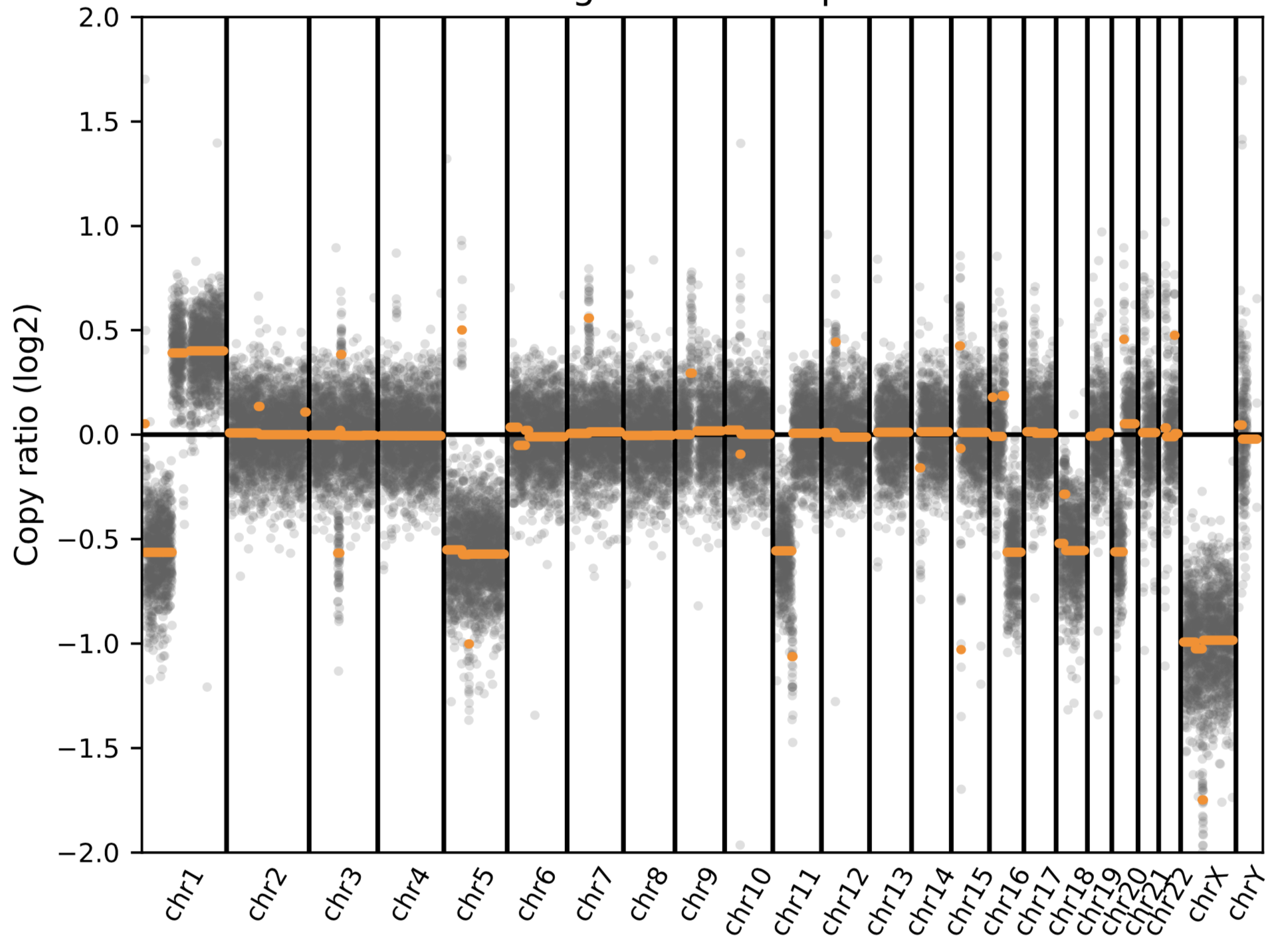 |
